# Association Between Glucagon-like Peptide-1 Receptor Agonist Use and Perioperative Aspiration: A Systematic Review and Meta-Analysis

**DOI:** 10.1101/2024.11.10.24317070

**Authors:** Jasmin Elkin, Siddharth Rele, Priya Sumithran, Michael Hii, Sharmala Thuraisingam, Tim Spelman, Tuong Phan, Peter Choong, Michelle Dowsey, Cade Shadbolt

## Abstract

**Background:** Glucagon-like peptide-1 receptor agonists (GLP-1 RAs) are known to slow gastric emptying, however the association between GLP-1 RA use and perioperative aspiration risk is not known. This systematic review and meta-analysis aimed to summarise the evidence on whether GLP-1 RA exposure is associated with (1) pulmonary aspiration in patients undergoing procedures requiring anaesthesia or sedation, or (2) increased residual gastric contents among fasted patients.

**Methods:** A search of MEDLINE, EMBASE, Web of Science, and Cochrane Central ClinicalTrials.gov and WHO ICTRP registries (updated 13 Jan 2025), and citation tracking of included studies was performed (14 Jan 2025). Studies assessing perioperative pulmonary aspiration or residual gastric contents among fasted patients who were using any form of GLP-1 RA were included. Data was extracted independently and in duplicate. Pooled odds ratios (ORs) were estimated for each outcome using random effect meta-analysis. Certainty of the evidence for each outcome was assessed using the Grading of Recommendations Assessment, Development, and Evaluation framework.

**Results:** Of 9,010 screened studies, 28 observational studies were included in the analysis. In a meta-analysis of 9 studies involving 304,060 individuals and 481 cases of aspiration, GLP-1RA exposure was not associated with pulmonary aspiration (OR, 1.04; 95% CI, 0.87-1.25, low certainty evidence). In a meta-analysis of 18 studies involving 165,522 individuals and 3,831 cases of residual gastric contents, GLP-1RA exposure was positively associated with residual gastric contents despite appropriate fasting (OR, 5.96; 95% CI, 3.96-8.98, low certainty evidence). In a meta-analysis of 5 studies involving 1,706 individuals and 208 cases of residual gastric contents, withholding at least one dose of GLP-1 RA prior to a procedure was associated with a lower odds of residual gastric contents (OR, 0.51; 95% CI, 0.33-0.81, very low certainty evidence). No studies measured the association between the time since last dose of GLP-1 RA and pulmonary aspiration.

**Conclusions:** Patients using GLP-1RAs are at heightened risk of presenting to surgery with residual gastric contents, though the available evidence does not indicate that this translates to an elevated risk of aspiration. Further research is needed to evaluate the risks and benefits of different strategies for managing these medications during the perioperative period.

## Introduction

Glucagon-like peptide-1 receptor agonists (GLP-1 RAs) are widely used for management of obesity and type 2 diabetes.^1^ GLP-1 RAs are known to slow the rate of postprandial gastric emptying,^2–4^ and there have been recent anecdotal accounts of patients who are taking these medications before surgery aspirating or having increased residual gastric contents despite appropriate fasting.^5–14^ This has led various professional organisations to acknowledge that patients taking these medications may be at a heightened risk of pulmonary aspiration or regurgitation during the perioperative period.^15–17^ In response, safety-related warnings regarding pulmonary aspiration risks in patients undergoing elective procedures were added to GLP-1 RA labels in November 2024.^18^

Currently, appropriate perioperative management strategies for patients using GLP-1 RAs remains uncertain. Several professional bodies have considered withholding GLP-1 RA medications before surgery, but emphasise that there is limited evidence available about effectiveness of this strategy.^16, 19–21^ Moreover, concerns have been raised about the safety of requiring an extended period of withdrawal, which may result in worsening glycaemic control among patients managing type 2 diabetes.^21–23^ Alternative strategies, including changing preoperative fasting guidance, using prokinetic agents, and implementing routine point-of-care ultrasound to mitigate risk have been considered, but their effectiveness also remains uncertain.^21, 24^

Previous systematic reviews have examined the risk of aspiration or residual gastric contents among GLP-1 RA users, however they have been limited to patients undergoing endoscopic procedures,^25–27^ or those temporarily exposed to GLP-1 Ras during the immediate perioperative period.^28^ To address uncertainty, this systematic review aimed to summarise the available evidence on whether GLP-1 RA exposure is associated with: (1) pulmonary aspiration in patients undergoing procedures requiring anaesthesia or sedation, or (2) increased residual gastric contents among fasted patients. This review also aimed to synthesise the evidence on whether withholding GLP-1 RA medications is associated with reductions in the risk of perioperative aspiration or the presence of residual gastric contents in fasted patients.

## Methods

We conducted a systematic review and meta-analysis in line with our prespecified protocol (PROSPERO: CRD42024532229) and reported according to the Preferred Reporting Items for Systematic Reviews and Meta-Analyses (PRISMA) statement^29^ and Meta-analyses Of Observational Studies in Epidemiology (MOOSE) reporting guidelines.^30^ Deviations and clarifications to the initial protocol are presented in the supplement (online Supporting Information Table S1). During the review process, discordance between reviewers were resolved through discussion, or via adjudication by an additional reviewer if consensus could not initially be achieved.

### Search strategy

We searched MEDLINE, EMBASE, Web of Science, and Cochrane Central on 19 March 2024 and completed an updated search on 13 Jan 2025 (online Supporting Information Methods S1). The search was limited to studies published from 2005 onwards, as this was the year of the first approval of a GLP1-RA by the Food and Drug Administration. The ClinicalTrials.gov and WHO ICTRP registries were searched on the same date for information on unpublished or ongoing studies. Forward and backwards citation tracking of included studies was last updated on 14 Jan 2025 using an automated online platform that has been described previously.^31^

### Selection criteria

Randomised controlled trials and observational studies were eligible for inclusion if they reported on the association between preoperative GLP-1 RA use and risk of: (1) pulmonary aspiration in fasted patients undergoing anaesthesia or procedural sedation; or (2) residual gastric contents in fasted patients undergoing procedures requiring anaesthesia or procedural sedation or in healthy volunteers that had fasted for at least 6 hours from solid foods and at least 2 hours for clear liquids, in accordance with standard preoperative fasting requirements.^32^

Studies were excluded if fasting duration was not reported, except for studies involving elective procedures where fasting was assumed to be a requirement. We also excluded studies where the GLP1-RA was commenced in the immediate perioperative period (i.e., within 14 days prior to the procedure or outcome measurement). Non-English studies and conference abstracts were not eligible for inclusion. When multiple studies were derived from overlapping samples, each study was included in the narrative summary though only the study with the lowest risk of bias across all 6 domains was included in pooled estimates. When the same study has been reported across multiple publications, only the most recent publication was included. Ongoing or completed study without published results were identified during screening, but not included in the quantitative synthesis. Studies were screened independently and in duplicate (by JE and SR) using the Covidence systematic review software.^33^ Reasons for excluded studies were reported and study authors were contacted to provide further information if eligibility was unclear.

### Outcome measures

Pulmonary aspiration included both direct measures of perioperative aspiration or postoperative respiratory complications that were directly attributed to aspiration (e.g. aspiration pneumonitis). Other respiratory complications that were not specifically attributed to perioperative aspiration did not meet this outcome definition. Residual gastric content included gastric residue identified via direct visualisation during endoscopy of the upper gastrointestinal tract or gastric contents identified via ultrasound. Individual outcome definitions as described by the study authors are presented in the supplement (online Supporting Information Table S2-S3).

### Data extraction and preparation

Key study characteristics and outcome data were extracted from each of the included studies, independently and in duplicate (by CS and JE) using a structured data extraction template. Adjusted odds ratios (OR) were the preferred effect estimates included in the meta-analyses of both outcomes. For estimates relating to aspiration, adjusted measures of relative risk (RR) were treated as adjusted ORs as odds approximates risk for low event rates.^34^ When adjusted estimates were not reported, unadjusted ORs were calculated from the reported number of aspiration events in patients with and without GLP-1 RA exposure. For studies with no events in at least one arm, we added a continuity correction value to both arms that was inversely proportional to the sample size of the other study arm.^34, 35^ For estimates relating to residual gastric contents, adjusted RRs were not treated as analogous to ORs, as this is a more common outcome. Study authors were contacted to provide adjusted ORs if multivariable analyses were reported alongside other measures of effect (online Supporting Information Table S4). When adjusted ORs could not be obtained, unadjusted ORs were calculated directly from the reported number of patients with residual gastric contents in exposed and unexposed groups.

### Risk of bias

Risk of bias assessment was conducted independently and in duplicate (by CS and JE) for each effect estimate rather than once per study.^36^ For studies examining the association between preoperative GLP-1 RA exposure as compared to no exposure and the risk of pulmonary aspiration or residual gastric contents, we used a modified Quality in Prognosis Studies (QUIPS) tool.^37,38^ For estimates of the association between withholding at least one dose of their GLP-1 RA and the study outcomes, we used the Risk Of Bias In Non-randomised Studies - of Interventions (ROBINS-I) tool. In line with ROBINS-I recommendations, studies judged to be at critical risk of bias were excluded from the meta-analyses.^39^ For each of the included studies, bias due to confounding was evaluated by consideration of adjustment for important variables, including age, sex, body mass index (BMI), diabetes, ASA-physical status classification system, medications associated with delaying gastric emptying, and comorbidity burden.

### Statistical analysis

Effect estimates for pulmonary aspiration and residual gastric contents were pooled separately. Pooled odds ratios and 95% confidence intervals were calculated via random effect meta-analysis using the restricted maximum likelihood (REML) heterogeneity variance estimator. ^40, 41^ Heterogeneity was assessed by estimating the I^2^ statistic. Funnel plots were assessed visually for evidence of small study bias, and the robustness of estimates to worst case publication bias was assessed by conducting meta-analyses including only non-affirmative studies.^42^ Overall certainty of the evidence for each pooled estimate was evaluated in line with the GRADE (Grading of Recommendations, Assessment, Development, and Evaluations) non-contextualised approach, which accounts for risk of bias, inconsistency, indirectness, imprecision, and publication bias of the included studies not considering the clinical context.^43^ All statistical analyses were performed using the *meta, metafor,* and *PublicationBias* packages in R Statistical Software (Version 4.3.2, R Core Team).^44, 45^

### Subgroup analyses

Two prespecified subgroups analyses were performed according to type of GLP-1 RA (i.e, study limited to once-weekly formulations vs study not limited to once-weekly formulations), and indication for GLP-1 RA (i.e, studies limited to patients with diabetes vs studies not limited to patients with diabetes). We performed a post-hoc subgroup analysis comparing studies limited to patients undergoing upper endoscopies to all other studies to examine whether the absence of association between GLP-1 RA exposure and pulmonary aspiration could be explained by altered intraoperative management following direct visualisation of residual gastric contents.

## Results

### Study Selection and Characteristics

The literature search identified 9,010 study records (Figure 1), from which 28 observational studies involving 466,373 patients were included in the meta-analysis (Table 1). Sixteen studies that were ongoing or completed without published results at the time of the final search are summarised separately in online Supporting Information Table S5.

**Figure 1.**
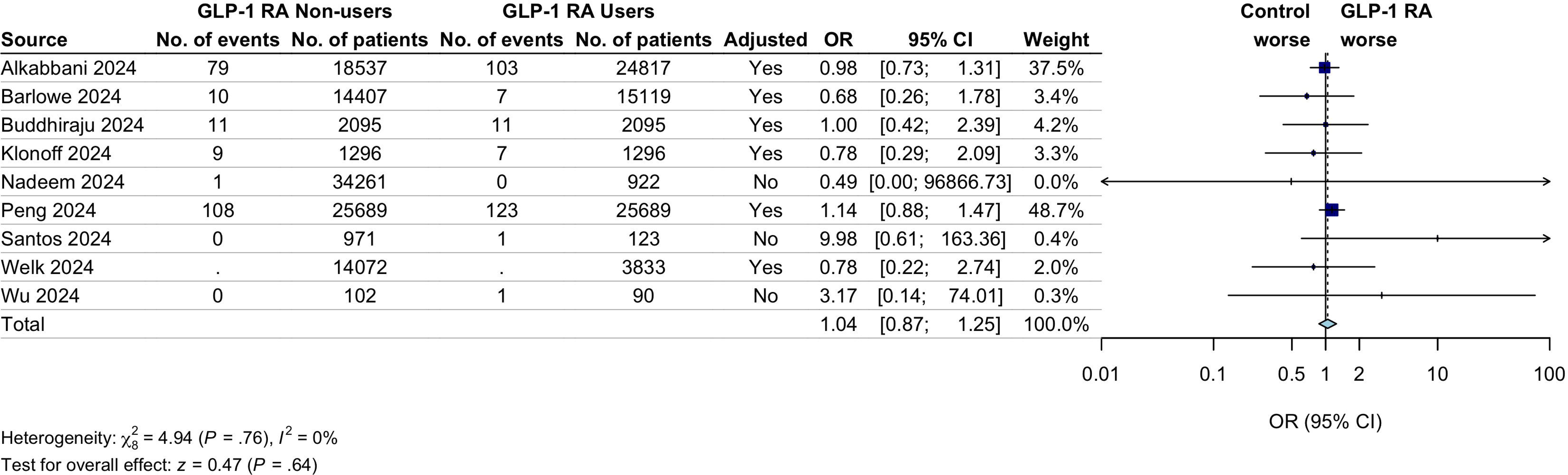
PRISMA Diagram. Abbreviations: GLP-1 RA, Glucagon-Like Peptide-1 Receptor Agonist

**Table 1.**
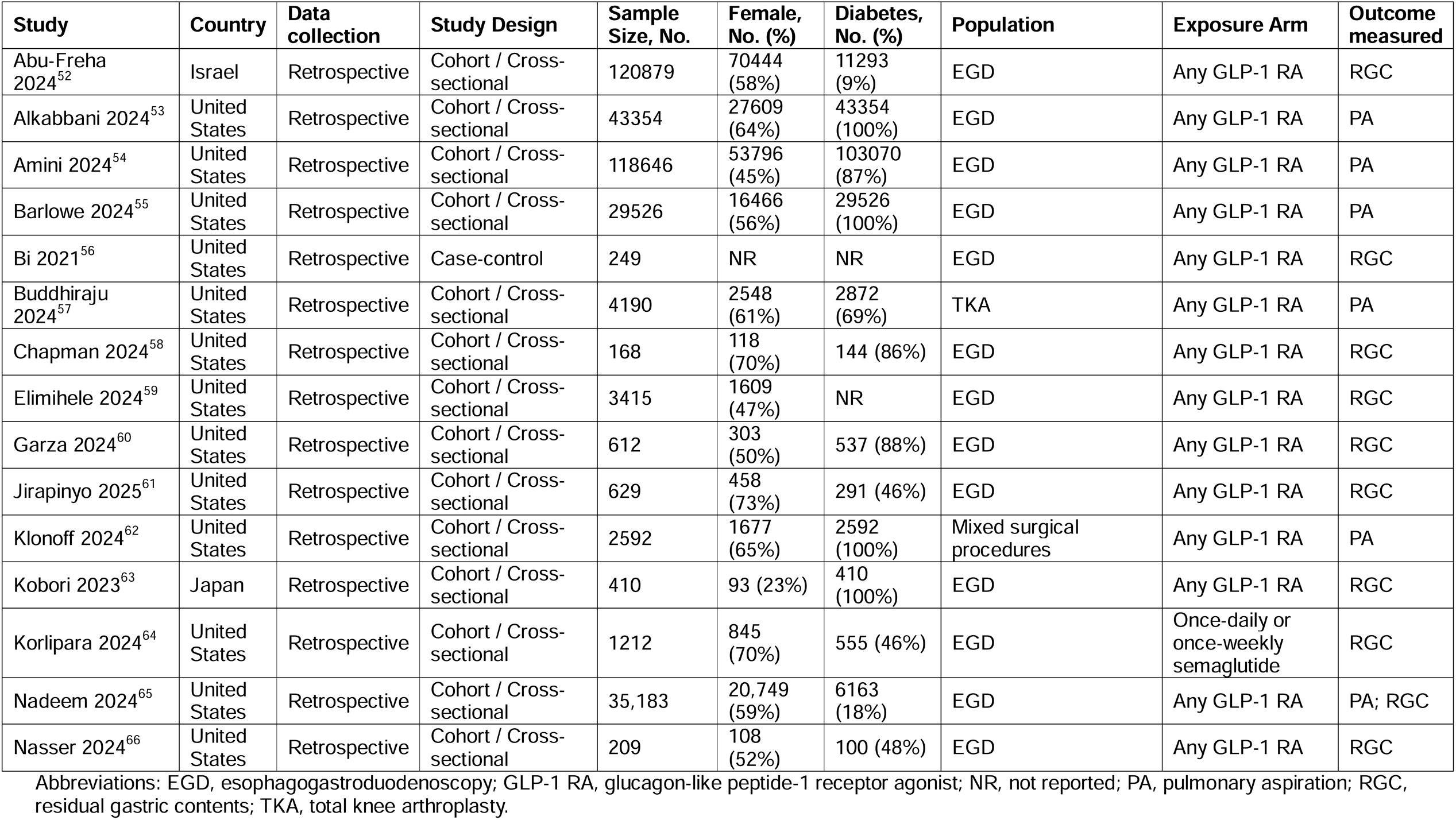
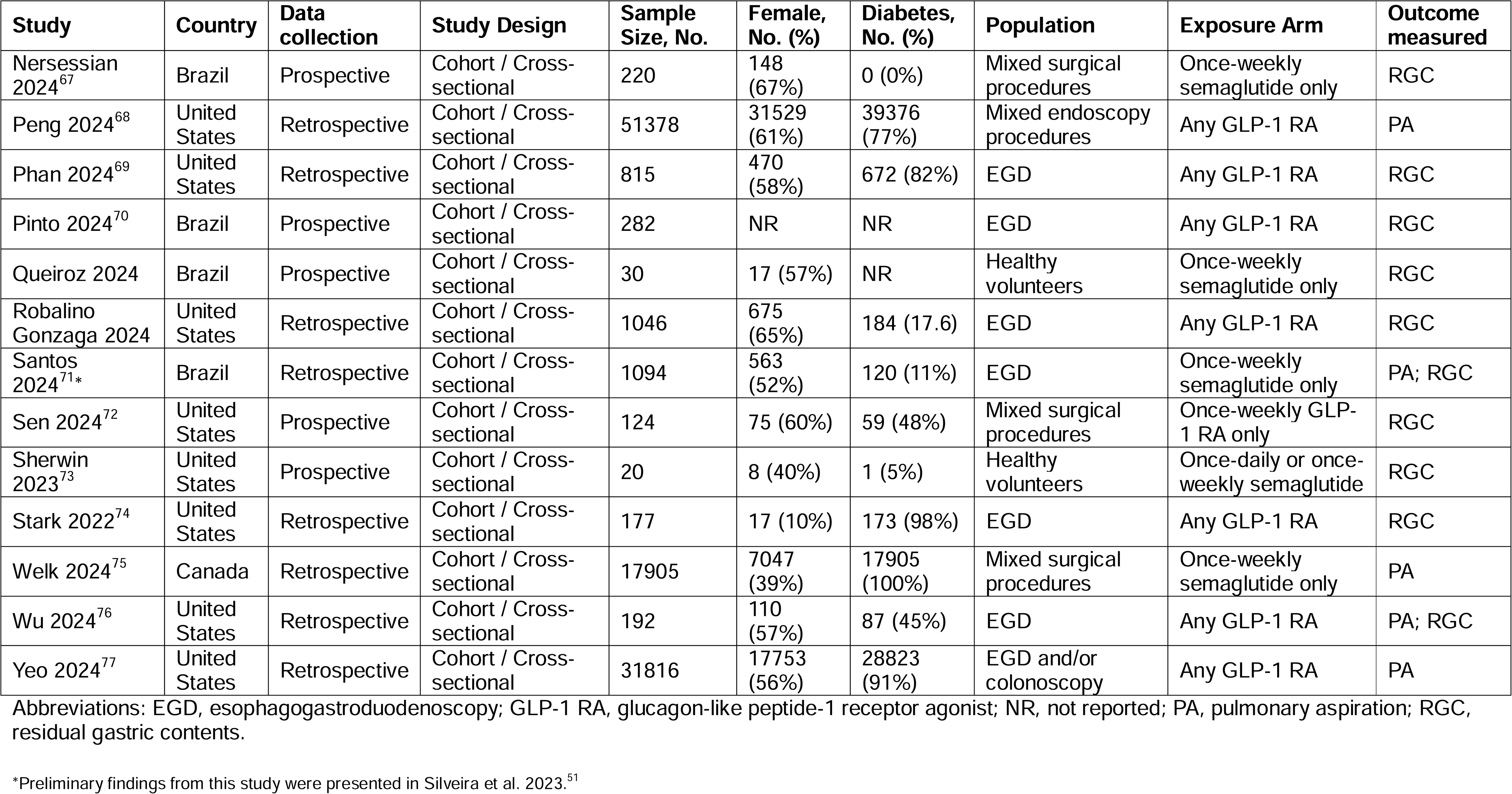
Characteristics of included studies.

### Pulmonary Aspiration

Eleven retrospective studies assessed pulmonary aspiration among 335,876 patients (Table 2). Of these studies, 6 (55%) included patients who underwent esophagogastroduodenoscopy (EGD), 1 (9%) included EGD and/or colonoscopies, 1 (9%) included mixed endoscopic procedures, 1 (9%) included total knee arthroplasty (TKA), and 2 (18%) involved other elective surgeries. Instances of pulmonary aspiration were identified from electronic medical records or administrative claims data (online Supporting Information Table S2). Three studies examined individuals undergoing similar procedures from the same administrative database. To avoid double counting of individuals,^46^ the study with the lowest risk of bias across all 6 domains was included in the meta-analysis.

**Table 2.**
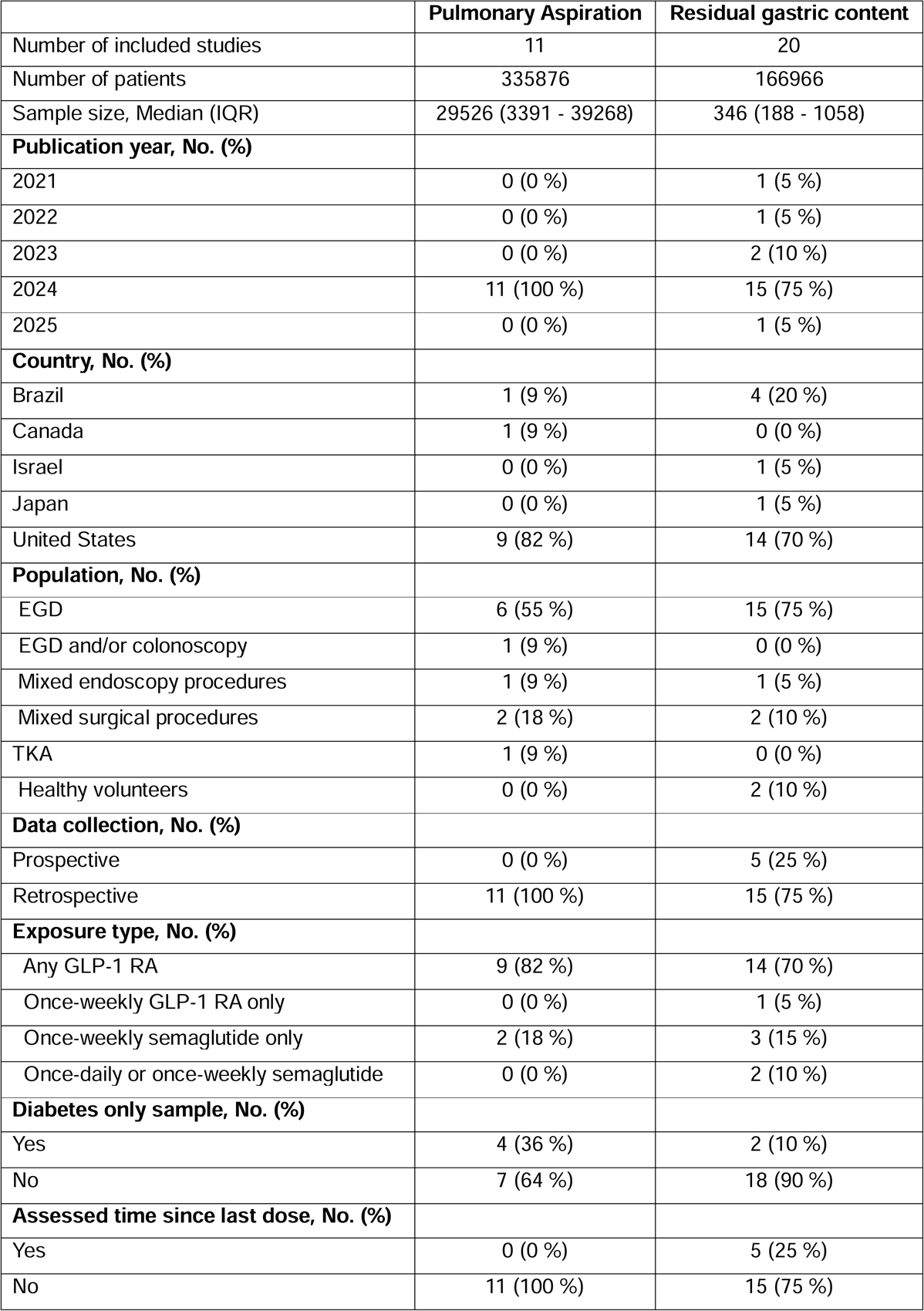
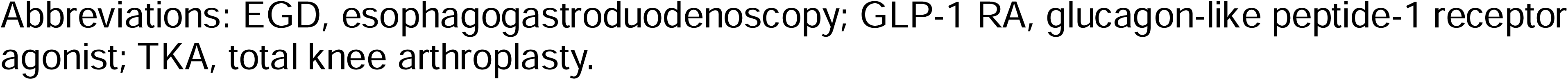
Characteristics of included studies by outcome.

Nine studies were included in the meta-analysis (Figure 2a). These studies examined 304,060 individuals, of which 481 cases of pulmonary aspiration were identified (Table 3). Preoperative exposure to GLP-1 RAs was not associated with pulmonary aspiration (OR, 1.04; 95% CI, 0.87-1.25, I^2^=0%) (Figure 2a). Evaluation of indirectness suggested potential concerns due to a large proportion of studies restricted to endoscopic procedures or patients with diabetes (Table 3). Visual inspection of funnel plot asymmetry did not indicate small study effects (online Supporting Information Figure S1). Sensitivity analysis restricted to non-affirmative studies was not performed due to an absence of affirmative studies. All subgroup analyses were consistent with the primary analysis (online Supporting Information Figure S2-S4). Overall risk of bias was moderate to high (online Supporting Information Table S6). For most studies, bias was largely attributed to incomplete adjustment for important prespecified confounders (online Supporting Information Table S7). Overall certainty in the evidence was low (Table 3).

**Figure 2a.**
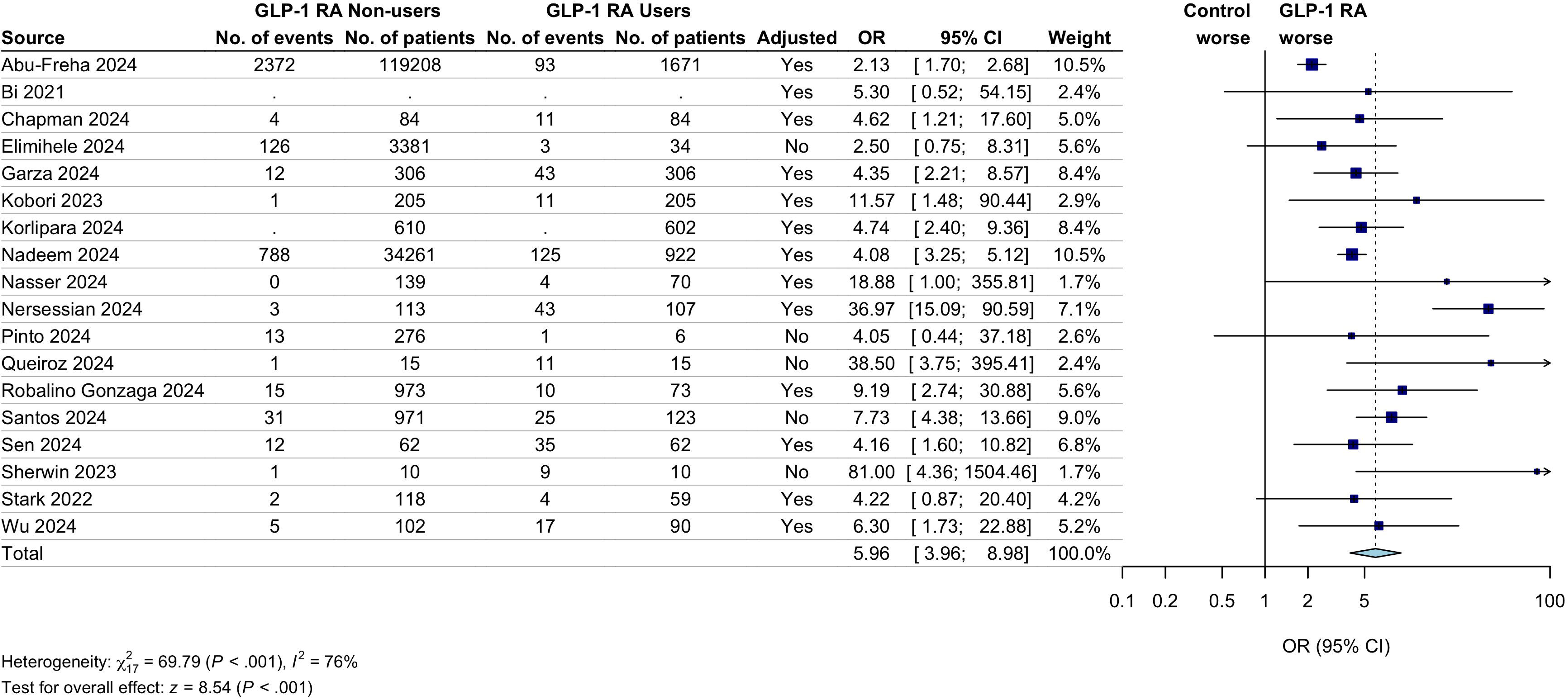
Forest plot pulmonary aspiration. Random-effects model method: restricted maximum likelihood heterogeneity variance estimator. The dark blue boxes represent individual study odds ratio, and the size of the boxes are proportional to study weight in the meta-analysis; the whiskers represent the confidence intervals; light blue diamond represents the overall pooled odds ratio and 95% CI; the dotted vertical line indicates the pooled OR. Event rates were not reported by Welk 2024. Abbreviations: GLP-1 RA, Glucagon-Like Peptide-1 Receptor Agonist.

**Table 3.**
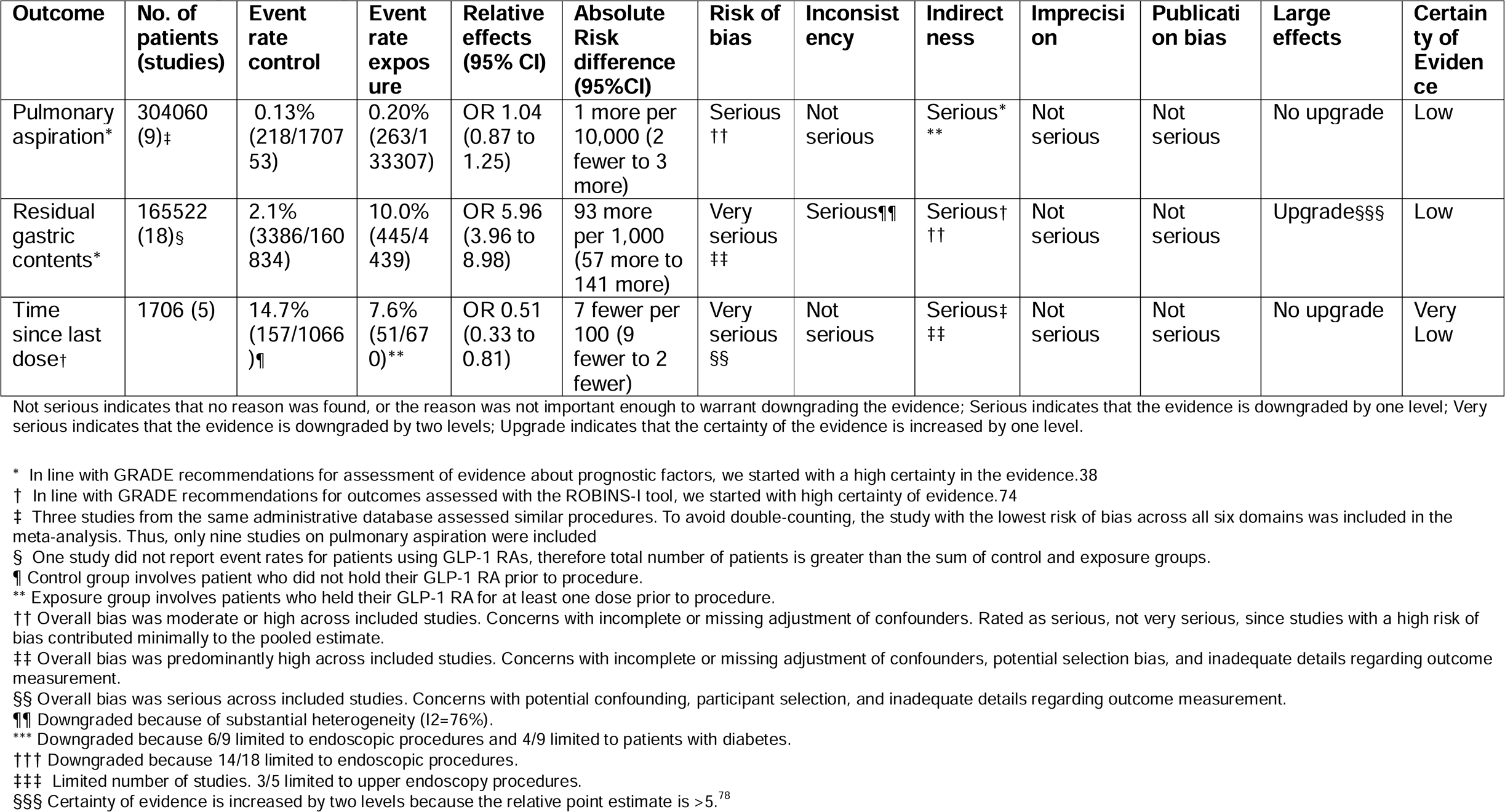
Summary of findings and GRADE assessment.

### Residual Gastric Contents

Twenty studies assessed residual contents (Table 2). Of these studies, 15 (75%) included patients who had undergone EGD procedures, 1 (5%) included various endoscopic procedures, 2 (10%) included other elective surgeries, and 2 (10%) included healthy volunteers. Residual gastric contents were identified via retrospective review of patient medical records for 15 studies involving endoscopic procedures, with the remaining 5 studies having measured residual gastric content prospectively using gastric ultrasound (online Supporting Information Table S3).

Eighteen studies were included in the meta-analysis (Figure 2b). These studies examined 165,522 individuals, of which 3,831 had residual gastric contents (Table 3). Preoperative GLP-1 RA exposure was associated with higher odds of residual gastric contents (OR, 5.96; 95% CI, 3.96-8.98; I^2^=76%) (Figure 2b), however this estimate was impacted by substantial heterogeneity. Although our certainty in evidence was increased due to the large effect, evaluation of indirectness suggested potential concerns due to a large proportion of studies restricted to endoscopic procedures (Table 3). Visual inspection of funnel plot asymmetry indicated that small study effects may have inflated this estimate (online Supporting Information Figure S5), though a sensitivity analysis restricted to non-affirmative studies indicated that our pooled estimate was robust to even worse-case publication bias (OR, 3.38; 95% CI, 1.49-7.68) (online Supporting Information Figure S6-S7). Subgroup analysis indicated that GLP-1 RA exposure had a stronger association with residual gastric contents in studies of patient who had not undergone an upper endoscopic procedure compared to studies of other populations (online Supporting Information Figure S8). Despite this, both subgroups showed a large effect in the same direction. No other subgroup effects were identified (online Supporting Information Figure S9 and S10). Overall risk of bias was predominantly high (online Supporting Information Table S8). For most studies, bias was largely attributed to high concerns with study participation, outcome measurement, and incomplete adjustment for important prespecified confounders (online Supporting Information Table S9). Overall certainty in the evidence was low (Table 3).

**Figure 2b.**
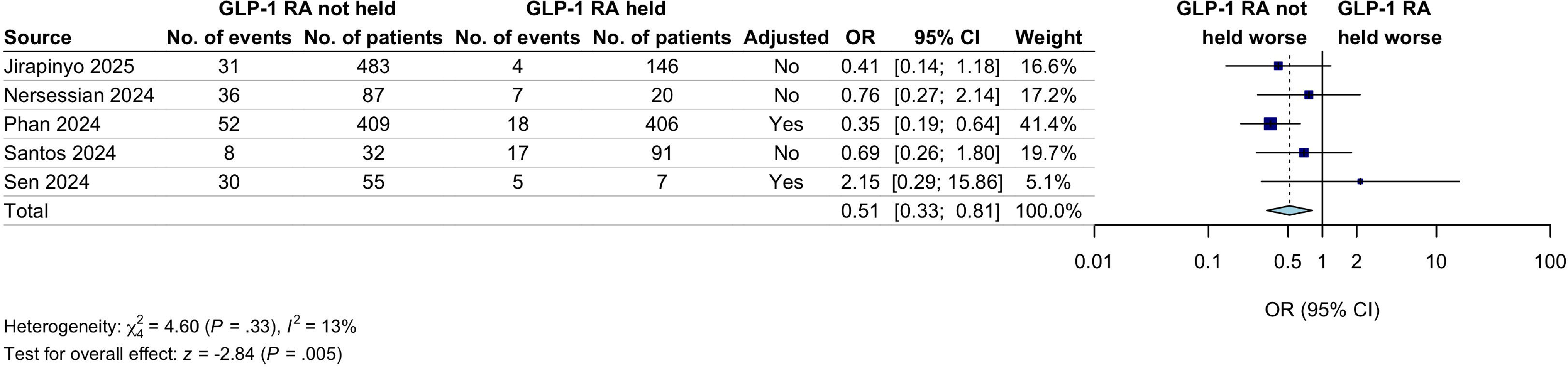
Forest plot residual gastric contents. Random-effects model method: restricted maximum likelihood heterogeneity variance estimator. The dark blue boxes represent individual study odds ratio, and the size of the boxes are proportional to study weight in the meta-analysis; the whiskers represent the confidence intervals; light blue diamond represents the overall pooled odds ratio and 95% CI; the dotted vertical line indicates the pooled OR. Event rates were not reported by Bi 2021 and Korlipara 2024. Abbreviations: GLP-1 RA, Glucagon-Like Peptide-1 Receptor Agonist.

### Association Between Time Since Last Dose and Residual Gastric Contents

Five observational studies assessed the association between the time since last dose of GLP-1RA medication and residual gastric contents (Table 2). These studies examined 1,706 patients who were using GLP-1 RA, of whom 208 had residual gastric contents (Table 3). Two of these studies examined a cohort using either weekly and daily dosed formulations, and the remaining studies were restricted to cohorts using weekly formulations (online Supporting Information Table S10).

All five studies were included in the meta-analysis (Figure2c). Withholding at least one dose of GLP-1 RA prior to a procedure was associated with a lower odds of residual gastric contents (OR, 0.51; 95% CI, 0.33-0.81; I^2^=13%). However, evaluation of indirectness suggested potential concerns due to a more than half of studies being limited to upper to EGD procedures. Visual inspection of funnel plot asymmetry indicated that small study effects may have inflated this estimate (online Supporting Information Figure S11). Sensitivity analysis restricted to non-affirmative studies indicated that the direction of the pooled estimate remained robust to even worse-case publication bias, though the wider confidence intervals from this sensitivity analysis included the null value (OR, 0.67; 95% CI, 0.38-1.18) (online Supporting Information Figure S12-S13). Subgroup analysis of GLP-1 RA indication was not performed due to an absence of studies restricted to patients with diabetes. No other subgroup effects were identified (online Supporting Information Figure S14 and S15). Overall risk of bias across studies analysed was serious (online Supporting Information Table S11). Key concerns were attributed to potential bias due to residual confounding, participant selection, and outcome measurement. Overall certainty in the evidence was very low (Table 3). No studies measured the association between the time since last dose of GLP-1 RA and pulmonary aspiration.

**Figure 2c.** Forest plot time since last dose. Random-effects model method: restricted maximum likelihood heterogeneity variance estimator. The dark blue boxes represent individual study odds ratio, and the size of the boxes are proportional to study weight in the meta-analysis; the whiskers represent the confidence intervals; light blue diamond represents the overall pooled odds ratio and 95% CI; the dotted vertical line indicates the pooled OR. Abbreviations: GLP-1 RA, Glucagon-Like Peptide-1 Receptor Agonist.

## Discussion

In this systematic review and meta-analysis, we assessed data from 28 observational studies involving 466,373 patients to evaluate the relationship between GLP-1 RA use and risk of perioperative aspiration among patients undergoing elective procedures. Our findings indicate that patients using GLP-1 RAs prior to an elective procedure are at a higher risk of presenting with residual gastric contents compared to those not using GLP-1 RAs. Despite this, the available evidence does not indicate that preoperative GLP-1 RA use is associated with risk of perioperative aspiration, and the relationship between withholding GLP-1 RAs before surgery and aspiration risk remains unstudied. Our analysis suggests that patients who withheld at least one dose of their GLP-1 RA before surgery were less likely to present with residual gastric contents, when compared to maintaining a regular dosing schedule, though this evidence was derived from five small observational studies that are inherently vulnerable to bias due to residual confounding. Importantly, available studies did not examine the potential risks of withholding these medications, such as glycaemic instability or abrupt increases in blood pressure.^47^ Given the absence of direct evidence from randomised controlled trials relating to such risks, and the small absolute risk of aspiration following appropriate fasting, it remains unclear if routinely withholding GLP-1 RA medications is warranted prior to elective procedures.

These findings may provide some degree of reassurance to both patients and clinicians about the risk of aspiration among patients using GLP-1 RAs prior to elective procedures. However, some caution is warranted, given low certainty in the available evidence. High rates of residual gastric contents among these patients may be a cause for concern, independent of any assumed association with aspiration risk. Gastric residue may complicate procedures that require clear stomach visualisation, such as gastroscopy, potentially leading to aborted procedures due to inadequate gastric clearance. Aborted procedures expose patients to unnecessary anaesthetic risks while also adversely affecting the efficient of healthcare resources.^48^ Consequently, these findings indicate that tailored guidance on the perioperative management of GLP-1 RAs may be warranted for procedures reliant on an empty stomach even in the absence of heightened aspiration risk.

This review highlights the need for targeted research exploring the risks and benefits of strategies for managing GLP-1 RA medications prior to elective procedures. While several observational studies were identified by our search of ongoing studies, it is unclear whether their findings from additional observational studies will add to the current evidence in a way that alters clinical practice. Importantly, two ongoing randomised controlled trials focused on preoperative management of GLP-1 RAs were identified. One assessing the effect of withholding GLP-1 RAs on aspiration and increased residual gastric content risk (NCT06533527), and another assessing the effectiveness of a 24-hour clear liquid diet for reducing residual gastric contents (NCT06654219). Such trials are urgently needed to assess the efficacy and safety of these approaches, and of alternative strategies, such as use of promotility agents, or assessing aspiration risk via preoperative gastric ultrasound.^24^ A rapid update to this review will be warranted once data is available from randomised controlled trials, as such evidence will be critical to informing recommendations presented in future clinical practice guidelines.

### Limitations

The findings of this review should be interpreted within the context of several limitations. First, there was heterogeneity in how outcomes were described across the included studies. This included differences in measurement methods, such as using endoscopy versus gastric ultrasound to assess gastric contents, and differences in outcome definitions, such as distinguishing between pulmonary aspiration and aspiration pneumonitis. Second, the available evidence is drawn largely from populations undergoing upper endoscopies, a procedure that is unique insofar as it allows for intraoperative management to be altered following direct visualisation of residual gastric contents. While this may limit the applicability of our findings to patients undergoing most types of elective procedures, the clinical interpretation of the findings were consistent across subgroup analyses of studies limited to patients undergoing upper endoscopies. Third, an absence of studies with a pre-registered protocol raises concerns about potential selective analysis and reporting, though the findings appeared to be largely robust to publication bias.

Fourth, none of the studies were judged to be a low risk of bias, which is reflected in the overall certainty of the evidence being low to very low. Finally, we excluded two potentially eligible studies that were not published in English.^49, 50^ It is unclear if this exclusion is likely to have altered our findings.

## Conclusion

This systematic review and meta-analysis found that patients using GLP-1 RAs are at a heightened risk of presenting for surgery with residual gastric contents, though this does not necessarily indicate an increased risk of aspiration. The available evidence to support withholding GLP-1 RAs before surgery is derived from a small number of observational studies and is impacted by a high degree of uncertainty. Given the uncertainty of evidence and absence of randomised controlled trials, ongoing research is needed to evaluate the risks and benefits of different strategies for managing these medications during the perioperative period.

## Data Availability

All data produced in the present study are available upon reasonable request to the authors

## Acknowledgements

None

## Details of authors’ contributions

JE and CS had full access to all the data in the study and take responsibility for the integrity of the data and the accuracy of the data analysis.

## Supporting Information captions

**Online Supporting Information Table S1**

Abbreviations: DPP-4, dipeptidyl peptidase-4; GLP-1 RA, glucagon-like peptide-1 receptor agonist; QUIPS, Quality in Prognosis Studies; RGC, residual gastric contents; RoB 2, Revised Cochrane risk-of-bias tool for randomised trials, ROBINS-E, Risk Of Bias In Non-randomised Studies – Of Exposure; ROBINS-I, Risk Of Bias In Non-randomised Studies - of Interventions (ROBINS-I).

*Preliminary findings from this study were presented in Silveira et al. 2023.^51^

**Online Supporting Information Table S2**

Abbreviations: EGD, Esophagogastroduodenoscopy; EMR, electronic medical record; ICD-9, International Classification of Diseases 9th Revision; ICD-10, International Classification of Diseases 10th Revision; ICD-10-CM, International Classification of Disease, Tenth Revision, and Clinical Modification; SNOMED, Systematized nomenclature of medicine; TKA, Total knee arthroplasty.

**Online Supporting Information Table S3**

Abbreviations: DGE, delayed gastric emptying; EGD, esophagogastroduodenoscopy; EMR, electronic medical record; GLP-1 RA, glucagon-like peptide-1 receptor agonist.

Abbreviations: EGD, esophagogastroduodenoscopy; EMR, electronic medical record; GLP-1 RA, glucagon-like peptide-1 receptor agonist; RGC, residual gastric contents.

**Online Supporting Information Table S4**

Abbreviations: GLP-1 RA, glucagon-like peptide-1 receptor agonist.

**Online Supporting Information Table S5**

Abbreviations: EGD, esophagogastroduodenoscopy; ERCP, endoscopic retrograde cholangiopancreatography; EUS, endoscopic ultrasound; GUS, gastric ultrasound; PA, pulmonary aspiration; RCT, Randomised controlled trial; RGC, residual gastric contents.

**Online Supporting Information Table S6**

Green indicates low risk of bias; Yellow indicates moderate risk of bias; Red indicates high risk of bias.

* Studies were rated as low risk of bias if all or most of the 6 domains were marked as low; Studies were rated as moderate risk of bias if most of the 6 domains were marked as moderate and none were marked as high; Studies were rated as high risk of bias if at least one domain was marked as high.

**Online Supporting Information Table S7**

Abbreviations: ASA-PS, American Society of Anesthesiologists Physical Status; BMI, Body mass index; CCB, Calcium channel blockers; CCI, Charlson comorbidity index; CKD, Chronic kidney disease; COPD, Chronic obstructive pulmonary disease; DPP-4, Dipeptidyl peptidase 4; GERD, Gastroesophageal reflux disease; GI, gastrointestinal; H2 blocker, Histamine type-2 receptor antagonists; MRA, Mineralocorticoid receptor antagonists; NA, Not applicable; NAFL, Non-alcoholic fatty liver disease; NASH, Non-alcoholic steatohepatitis; PPI, Proton pump inhibitor.

* Studies marked as NA restricted their inclusion criteria to patients with diabetes.

**Online Supporting Information Table S8**

Green indicates low risk of bias; Yellow indicates moderate risk of bias; Red indicates high risk of bias. Abbreviations: NA, not applicable.

* Studies marked as NA due to acute presentation with residual gastric contents, therefore assessment of study attrition is not relevant.

† Studies were rated as low risk of bias if all or most of the 6 domains were marked as low; Studies were rated as moderate risk of bias if most of the 6 domains were marked as moderate and none were marked as high; Studies were rated as high risk of bias if at least one domain was marked as high.

**Online Supporting Information Table S9**

Abbreviations: ASA-PS, American Society of Anesthesiologists physical status; BMI, Body mass index; CKD, Chronic kidney disease; EGD, Esophagogastroduodenoscopy; GLP-1 RA, Glucagon-like peptide-1 receptor agonist; HbA1C, Haemoglobin A1C; NA, Not applicable; NR, Not reported; PPI, Proton pump inhibitor.

* Studies marked as NA restricted their inclusion criteria to patients with diabetes.

† Variables adjusted for in the multivariate logistic regression were not clearly reported.

‡ Propensity score matching was performed using the variables listed in the table. Variables adjusted for in the multivariate logistic regression were not clearly reported.

**Online Supporting Information Table S10**

Abbreviations: CI, confidence interval; GLP-1 RA, glucagon-like peptide-1 receptor agonist; NR, not reported; OR, odds ratio; PR, prevalence ratio.

* Indicates patients who had their last GLP-1 RA dose either > 7 days (weekly doses) or >1 day (daily doses) prior to measurement of residual gastric contents.

† Indicates patients who had their last GLP-1 RA dose ≤ 7 days (weekly doses) or ≤ 1 day (single doses) prior to measurement of residual gastric contents.

‡ Patients who did not hold their GLP-1 RA prior to procedure were meta-analysed as the control group. Where appropriate, we took the inverse of published estimates to ensure consistency in the direction of the pooled effect estimates.

§ Study did not report effect estimates. Unadjusted odds ratio was calculated using the unadjusted event rates reported in the manuscript.

**Online Supporting Information Table S11**

Green indicates low risk of bias; Yellow indicates moderate risk of bias; Orange indicates serious risk of bias; Red indicates critical risk of bias; Grey indicates no information to form a judgement about risk of bias. Abbreviations: RGC, residual gastric contents.

* Studies were rated as low risk of bias if all domains were marked as low; Studies were rated as moderate risk of bias if all domains are either low or moderate; Studies were rated as serious risk of bias if at least one domain was marked as serious, but not at critical risk of bias in any domain; Studies were rated as critical risk of bias if at least one domain was marked as critical.

**Online Supporting Information Figure S1**

Funnel plot for pulmonary aspiration outcome. Grey dots indicate point estimate and standard error for each of the included studies assessing pulmonary aspiration.

**Online Supporting Information Figure S2**

Subgroup analysis for pulmonary aspiration outcome comparing studies of patients with diabetes only to studies of patients with diabetes or another indication. Random-effects model method: restricted maximum likelihood heterogeneity variance estimator. The dark blue boxes represent individual study odds ratio, and the size of the boxes are proportional to study weight in the meta-analysis; the whiskers represent the confidence intervals; light blue diamond represents the overall pooled odds ratio and 95% CI; the dotted vertical line indicates the pooled OR. Abbreviations: GLP-1 RA, Glucagon-Like Peptide-1 Receptor Agonist.

**Online Supporting Information Figure S3**

Subgroup analysis for pulmonary aspiration outcome comparing studies of upper endoscopies to all other studies. Random-effects model method: restricted maximum likelihood heterogeneity variance estimator. The dark blue boxes represent individual study odds ratio, and the size of the boxes are proportional to study weight in the meta-analysis; the whiskers represent the confidence intervals; light blue diamond represents the overall pooled odds ratio and 95% CI; the dotted vertical line indicates the pooled OR. Abbreviations: GLP-1 RA, Glucagon-Like Peptide-1 Receptor Agonist.

**Online Supporting Information Figure S4**

Subgroup analysis for pulmonary aspiration outcome comparing studies limited to once-weekly formulations vs study not limited to once-weekly formulations. Random-effects model method: restricted maximum likelihood heterogeneity variance estimator. The dark blue boxes represent individual study odds ratio, and the size of the boxes are proportional to study weight in the meta-analysis; the whiskers represent the confidence intervals; light blue diamond represents the overall pooled odds ratio and 95% CI; the dotted vertical line indicates the pooled OR. Abbreviations: GLP-1 RA, Glucagon-Like Peptide-1 Receptor Agonist.

**Online Supporting Information Figure S5**

Funnel plot for residual gastric contents outcome. Grey dots indicate point estimate and standard error for each of the included studies assessing pulmonary aspiration.

**Online Supporting Information Figure S6**

Significance plot for residual gastric contents outcome. Grey dot indicates non-affirmative studies; orange dot indicates affirmative studies; black diamond indicates estimate from meta-analysis for all studies; grey diamond indicates sensitivity analysis of non-affirmative studies.

**Online Supporting Information Figure S7**

Sensitivity analysis of non-affirmative studies for residual gastric contents outcome. Random-effects model with restricted maximum likelihood heterogeneity variance estimator. The dark blue boxes represent individual study odds ratio, and the size of the boxes are proportional to study weight in the meta-analysis; the whiskers represent the confidence intervals; light blue diamond represents the overall pooled odds ratio and 95% CI; the dotted vertical line indicates the pooled OR. Adjusted indicates adjusted ORs. Abbreviations: GLP-1 RA, Glucagon-Like Peptide-1 Receptor Agonist.

**Online Supporting Information Figure S8**

Subgroup analysis for residual gastric contents outcome comparing studies of upper endoscopies to all other studies. Random-effects model method: restricted maximum likelihood heterogeneity variance estimator. The dark blue boxes represent individual study odds ratio, and the size of the boxes are proportional to study weight in the meta-analysis; the whiskers represent the confidence intervals; light blue diamond represents the overall pooled odds ratio and 95% CI; the dotted vertical line indicates the pooled OR. Abbreviations: GLP-1 RA, Glucagon-Like Peptide-1 Receptor Agonist.

**Online Supporting Information Figure S9**

Subgroup analysis for residual gastric contents outcome comparing studies of patients with diabetes only to studies of patients with diabetes or another indication. Random-effects model method: restricted maximum likelihood heterogeneity variance estimator. The dark blue boxes represent individual study odds ratio, and the size of the boxes are proportional to study weight in the meta-analysis; the whiskers represent the confidence intervals; light blue diamond represents the overall pooled odds ratio and 95% CI; the dotted vertical line indicates the pooled OR. Abbreviations: GLP-1 RA, Glucagon-Like Peptide-1 Receptor Agonist.

**Online Supporting Information Figure S10**

Subgroup analysis for residual gastric contents outcome comparing studies limited to once-weekly formulations vs study not limited to once-weekly formulations. Random-effects model method: restricted maximum likelihood heterogeneity variance estimator. The dark blue boxes represent individual study odds ratio, and the size of the boxes are proportional to study weight in the meta-analysis; the whiskers represent the confidence intervals; light blue diamond represents the overall pooled odds ratio and 95% CI; the dotted vertical line indicates the pooled OR. Abbreviations: GLP-1 RA, Glucagon-Like Peptide-1 Receptor Agonist.

**Online Supporting Information Figure S11**

Funnel plot for studies assessing withholding at least one GLP-1 RA dose prior to a procedure on residual gastric contents. Grey dots indicate point estimate and standard error for each of the included studies assessing pulmonary aspiration.

**Online Supporting Information Figure S12**

Significance plot for studies assessing withholding at least one GLP-1 RA dose prior to a procedure on residual gastric contents. Grey dot indicates non-affirmative studies; orange dot indicates affirmative studies; black diamond indicates estimate from meta-analysis for all studies; grey diamond indicates sensitivity analysis of non-affirmative studies.

**Online Supporting Information Figure S13**

Sensitivity analysis of non-affirmative studies for studies assessing withholding at least one GLP-1 RA dose prior to a procedure on residual gastric contents. Random-effects model with restricted maximum likelihood heterogeneity variance estimator.

The dark blue boxes represent individual study odds ratio, and the size of the boxes are proportional to study weight in the meta-analysis; the whiskers represent the confidence intervals; light blue diamond represents the overall pooled odds ratio and 95% CI; the dotted vertical line indicates the pooled OR. Adjusted indicates adjusted ORs. Abbreviations: GLP-1 RA, Glucagon-Like Peptide-1 Receptor Agonist.

**Online Supporting Information Figure S14**

Subgroup analysis for studies assessing withholding at least one GLP-1 RA dose prior to a procedure on residual gastric contents, comparing studies of upper endoscopies to all other studies. Random-effects model method: restricted maximum likelihood heterogeneity variance estimator. The dark blue boxes represent individual study odds ratio, and the size of the boxes are proportional to study weight in the meta-analysis; the whiskers represent the confidence intervals; light blue diamond represents the overall pooled odds ratio and 95% CI; the dotted vertical line indicates the pooled OR. Abbreviations: GLP-1 RA, Glucagon-Like Peptide-1 Receptor Agonist.

**Online Supporting Information Figure S15**

Subgroup analysis for studies assessing withholding at least one GLP-1 RA dose prior to a procedure on residual gastric contents, comparing studies limited to once-weekly formulations vs study not limited to once-weekly formulations. Random-effects model method: restricted maximum likelihood heterogeneity variance estimator. The dark blue boxes represent individual study odds ratio, and the size of the boxes are proportional to study weight in the meta-analysis; the whiskers represent the confidence intervals; light blue diamond represents the overall pooled odds ratio and 95% CI; the dotted vertical line indicates the pooled OR. Abbreviations: GLP-1 RA, Glucagon-Like Peptide-1 Receptor Agonist.

