## Supplementary material for "Association Between Glucagon-like Peptide-1 Receptor Agonist Use and Perioperative Aspiration: A Systematic Review and Meta-Analysis": online Supporting Information

### **Table S1. Deviations and clarifications to the initial protocol**

| **Protocol description** | **Deviation or Clarification** |
| --- | --- |
| “For our primary questions, our comparison of interest will be no GLP1-RA medication use prior to surgery/fasting.” | The study protocol did not adequately specify how comparators would be selected when multiple comparisons were available. Barlowe et al. reported comparisons between GLP-1 RA and DPP-4 Inhibitor users, and GLP-1 RA and opioid users.^1^ DPP-4 Inhibitor users were selected as the comparator in our analysis, as Barlowe et al. selected them as a negative control because DPP-4 Inhibitors have no known association with gastric emptying.^1^ |
| “Risk of bias will be assessed independently and in duplicate using the RoB 2 and ROBINS-E tools.” | During piloting of the ROBINS-E tool, it became evident that it was not appropriate for the questions assessed in this review. The association between pre-existing GLP-1 RA use and aspiration/RGC could not be formulated into a definitive causal question. As such, estimates relating to these associations were deemed to be prognostic rather than causal, and the QUIPs tool was deemed the most appropriate tool for assessing risk of bias. The prognostic question of whether patients using GLP-1 RA medications are at a higher risk of aspirating or presenting with increased residual gastric contents than the general population is of relevance to patients and clinicians.  In contrast, when assessing the association between holding vs not holding at least one dose of GLP-1 RA on residual gastric contents, this could be straightforwardly formulated as a causal question. As withholding these medications was deemed to be an intervention rather than an exposure, the ROBINS-I tool was used for evaluating bias for these effect estimates. |
| “This review aims to address the following secondary question: Is the association between GLP-1 RA exposure and aspiration risk or residual gastric contents/volume modified by the time since last exposure or duration of fasting?” | We did not specify how the time since last dose of GLP-1 RA would be operationalised in our analysis. When this became apparent during the review process, we decided to assess this association using the binary exposure variable reported by Phan et al.^2^ This approach was thought to be more relevant to the ongoing debate regarding withholding GLP-1 RAs prior to surgery, than assessing this using a continuous variable (e.g., days since last dose). |
| “If sufficient data is available, the following subgroups will be examined via meta-regression and/or subgroup specific meta-analyses.  - Type of GLP-1 RA (i.e. long vs short acting)  - Indication for GLP-1 RA (e.g. obesity, diabetes).” | For the subgroup analysis related to GLP-1 RA type we examined studies limited to patients using once-weekly GLP-1 RAs only, compared to those which included both once-weekly or once-daily formulations. No studies restricted their analysis to once-daily formulations. We also conducted a subgroup analysis of studies limited to patients with diabetes compared to studies not limited to patients with diabetes. As only one study was restricted patients without diabetes, a separate subgroup analysis of populations without diabetes was not conducted.    In addition, we conducted a post-hoc subgroup analysis on procedure type (i.e., studies limited to  patients undergoing upper endoscopic procedures versus all other studies), to examine whether the absence of association between GLP-1 RA exposure and pulmonary aspiration could be explained by altered intraoperative management following direct visualisation of residual gastric contents. |

Abbreviations: DPP-4, dipeptidyl peptidase-4; GLP-1 RA, glucagon-like peptide-1 receptor agonist**;** QUIPS, Quality in Prognosis Studies; RGC, residual gastric contents; RoB 2, Revised Cochrane risk-of-bias tool for randomised trials, ROBINS-E, Risk Of Bias In Non-randomised Studies – Of Exposure; ROBINS-I, Risk Of Bias In Non-randomised Studies - of Interventions (ROBINS-I

### **Methods S1. Search Strategies**

All search strategies were constructed by members of the review team (JE, CS, SR), which included members who have extensive experience with formulating systematic review searches (CS, SR). The search was conducted by the first author (JE).

| **Ovid Embase Classic+Embase** | | | | |
| --- | --- | --- | --- | --- |
| **Search Number** | **Search Strategy** | **Initial Search (19 Mar 2024)** | **Updated Search (21 Oct 2024)** | **Updated Search (13 Jan 2025)** |
| 1 | exp exendin 4/ or exp liraglutide/ or exp albiglutide/ or exp dulaglutide/ or exp lixisenatide/ or exp semaglutide/ or exp tirzepatide/ or exp glucagon like peptide/ or exp glucagon like peptide 1 receptor agonist/ | 55062 | 59798 | 61615 |
| 2 | (liraglutide or semaglutide or tirzepatide or exenatide or beinaglutide or dulaglutide or albiglutide or lixisenatide or loxenatide or efpeglenatide or polyethylene glycol loxenatide or mazdutide or orforglipron or retatrutide or PEG-exenatide or PEG loxenatide or pegloxenatide or LAPS-exendin or langlenatide or LAPS-exendin 4 analogue or LAPSCA-exendin4 or LAPS-exendin4 or LAPSEXD4 or exendin$4).ti,ab,kw. | 18346 | 19980 | 20631 |
| 3 | (Ozempic or Rybelsus or Wegovy or Victoza or Saxenda or Mounjaro or CagriSema or Byetta or Bydureon or Bydureon BCise or Presendin or Adlyxin or Lyxumia or Tanzeum or Eperzan or Syncria or Albugon or Trulicity or Zepbound or Soliqua or Xultophy).ti,ab,kw. | 655 | 776 | 830 |
| 4 | (GLP1* or GLP-1* or glucagon-like peptide* or incretin*).ti,ab,kw. | 40278 | 43292 | 44555 |
| 5 | 1 or 2 or 3 or 4 | 62545 | 67660 | 69591 |
| 6 | food aspiration/ or aspiration/ or pulmonary aspiration/ or aspiration pneumonia/ or liquid aspiration/ | 68978 | 70485 | 71026 |
| 7 | (aspirat* or regurgitat*).ti,ab,kw. | 282567 | 291395 | 295192 |
| 8 | stomach content/ or stomach emptying/ | 28906 | 29651 | 29979 |
| 9 | ((gastric or stomach or gastrointestinal) adj2 (content* or volume* or residual or residue or empty*)).ti,ab,kw. | 37910 | 38892 | 39301 |
| 10 | esophagogastroduodenoscopy/ or endoscopy/ or "point of care ultrasound"/ or ultrasound therapy/ or exp gastroscopy/ | 200330 | 209528 | 213813 |
| 11 | (ultrasound or sonograph* or ultrasonograph* or endoscop* or esophagogastroduodenoscop* or oesophagogastroduodenoscop* or gastroscop*).ti,ab,kw. | 1085342 | 1126384 | 1141426 |
| 12 | 6 or 7 or 8 or 9 or 10 or 11 | 1429638 | 1481748 | 1501325 |
| 13 | 5 and 12 | 3671 | 4017 | 4209 |
| 14 | exp animals/ not exp humans/ | 6015059 | 6121684 | 6161682 |
| 15 | 13 not 14 | 3328 | 3665 | 3852 |
| 16 | limit 15 to yr="2005 -Current" | 3120 | 3457 | 3643 |
| 17 | limit 16 to dc=20240319-20241021 | NA | 347 | NA |
| 17 | limit 16 to dc=20241021-20250113 | NA | NA | 208 |

| **Ovid MEDLINE(R** | | | | |
| --- | --- | --- | --- | --- |
| **Search Number** | **Search Strategy** | **Initial Search (19 Mar 2024)** | **Updated Search (21 Oct 2024)** | **Updated Search (13 Jan 2025)** |
| 1 | Exenatide/ or Liraglutide/ or glucagon-like peptide receptors/ or glucagon-like peptide-1 receptor/ or Glucagon-Like Peptide 1/ | 15822 | 16773 | 17090 |
| 2 | (liraglutide or semaglutide or tirzepatide or exenatide or beinaglutide or dulaglutide or albiglutide or lixisenatide or loxenatide or efpeglenatide or polyethylene glycol loxenatide or mazdutide or orforglipron or retatrutide or PEG-exenatide or PEG loxenatide or pegloxenatide or LAPS-exendin or langlenatide or LAPS-exendin 4 analogue or LAPSCA-exendin4 or LAPS-exendin4 or LAPSEXD4 or exendin$4).ti,ab,kw. | 9936 | 10986 | 11398 |
| 3 | (Ozempic or Rybelsus or Wegovy or Victoza or Saxenda or Mounjaro or CagriSema or Byetta or Bydureon or Bydureon BCise or Presendin or Adlyxin or Lyxumia or Tanzeum or Eperzan or Syncria or Albugon or Trulicity or Zepbound or Soliqua or Xultophy).ti,ab,kw. | 346 | 415 | 440 |
| 4 | (GLP1* or GLP-1* or glucagon-like peptide* or incretin*).ti,ab,kw. | 25281 | 27322 | 28201 |
| 5 | 1 or 2 or 3 or 4 | 29228 | 31680 | 32710 |
| 6 | exp Respiratory Aspiration/ or Pneumonia, Aspiration/ | 7412 | 7523 | 7568 |
| 7 | (aspirat* or regurgitat*).ti,ab,kw. | 176416 | 181111 | 183104 |
| 8 | gastrointestinal content/ or gastric emptying/ | 14857 | 15016 | 15075 |
| 9 | ((gastric or stomach or gastrointestinal) adj2 (content* or volume* or residual or residue or empty*)).ti,ab,kw. | 24404 | 24974 | 25200 |
| 10 | Endoscopy, Digestive System/ or Endoscopy/ or Gastroscopy/ or Ultrasonography/ | 288580 | 292877 | 294644 |
| 11 | (ultrasound or sonograph* or ultrasonograph* or endoscop* or esophagogastroduodenoscop* or oesophagogastroduodenoscop* or gastroscop*).ti,ab,kw. | 697519 | 721582 | 731480 |
| 12 | 6 or 7 or 8 or 9 or 10 or 11 | 963150 | 991688 | 1003411 |
| 13 | 5 and 12 | 1495 | 1627 | 1688 |
| 14 | exp animals/ not exp humans/ | 5203854 | 5268607 | 5297866 |
| 15 | 13 not 14 | 1322 | 1446 | 1505 |
| 16 | limit 15 to yr="2005 -Current" | 1154 | 1278 | 1337 |
| 17 | 16 and (202403* or 202404* or 202405* or 202406* or 202407* or 202408* or 202409* or 202410*).dt,ez,da. | NA | 148 | NA |
| 17 | 16 and (202410* or 202411* or 202412* or 202501*).dt,ez,da. | NA | NA | 93 |

| **Web of Science** | | | | |
| --- | --- | --- | --- | --- |
| **Search Number** | **Search Strategy** | **Initial Search (19 Mar 2024)** | **Updated Search (21 Oct 2024)** | **Updated Search (13 Jan 2025)** |
| 1 | TS=(liraglutide or semaglutide or tirzepatide or exenatide or beinaglutide or dulaglutide or albiglutide or lixisenatide or loxenatide or efpeglenatide or polyethylene glycol loxenatide or mazdutide or orforglipron or retatrutide or PEG-exenatide or PEG loxenatide or pegloxenatide or LAPS-Exendin or langlenatide or LAPS-exendin 4 analogue or LAPSCA-Exendin4 or LAPS-Exendin4 or LAPSEXD4 or exendin$4) | 13762 | 15322 | 15989 |
| 2 | TS=(Ozempic or Rybelsus or Wegovy or Victoza or Saxenda or Mounjaro or CagriSema or Byetta or Bydureon or Bydureon BCise or Presendin or Adlyxin or Lyxumia or Tanzeum or Eperzan or Syncria or Albugon or Trulicity or Zepbound or Soliqua or Xultophy) | 371 | 445 | 469 |
| 3 | TS=(GLP1* or GLP-1* or glucagon-like peptide* or incretin*) | 36593 | 39408 | 40761 |
| 4 | #1 OR #2 OR #3 | 42543 | 46017 | 47693 |
| 5 | TS=(aspirat* or regurgitat*) | 230766 | 247237 | 250244 |
| 6 | TS=((gastric or stomach or gastrointestinal) NEAR/2 (content* or volume* or residual or residue or empty*)) | 33693 | 38324 | 38677 |
| 7 | TS=(ultrasound or sonograph* or ultrasonograph* or endoscop* or esophagogastroduodenoscop* or oesophagogastroduodenoscop* or gastroscop*) | 871043 | 917343 | 930519 |
| 8 | #5 OR #6 OR #7 | 1095848 | 1160750 | 1176730 |
| 9 | #8 AND #4 | 1722 | 1898 | 2006 |
| 10 | PY=(2005-2024)* | 53593813 | 55914226 | 56810681 |
| 11 | #9 AND #10 | 1527 | 1700 | 1808 |
| 12 | #11 AND (LD=(2024-03-19/2024-10-21)) | NA | 173 | NA |
| 12 | #11 AND (LD=(2024-10-21/2025-01-13)) | NA | NA | 110 |

*Updated to PY=(2005-2024) for updated search on 13 Jan 2025

| **Cochrane CENTRAL** | | | | |
| --- | --- | --- | --- | --- |
| **Search Number** | **Search Strategy** | **Initial Search (19 Mar 2024)** | **Updated Search (21 Oct 2024)** | **Updated Search (13 Jan 2025)** |
| 1 | MeSH descriptor: [Glucagon-Like Peptide 1] explode all trees | 2402 | 2458 | 2489 |
| 2 | (liraglutide or semaglutide or tirzepatide or exenatide or beinaglutide or dulaglutide or albiglutide or lixisenatide or loxenatide or efpeglenatide or polyethylene glycol loxenatide or mazdutide or orforglipron or retatrutide or PEG-exenatide or PEG loxenatide or pegloxenatide or LAPS-exendin or langlenatide or LAPS-exendin 4 analogue or LAPSCA-exendin4 or LAPS-exendin4 or LAPSEXD4 or exendin$4):ti,ab,kw (Word variations have been searched) | 5707 | 6127 | 6301 |
| 3 | (Ozempic or Rybelsus or Wegovy or Victoza or Saxenda or Mounjaro or CagriSema or Byetta or Bydureon or Bydureon BCise or Presendin or Adlyxin or Lyxumia or Tanzeum or Eperzan or Syncria or Albugon or Trulicity or Zepbound or Soliqua or Xultophy):ti,ab,kw (Word variations have been searched) | 462 | 503 | 525 |
| 4 | (GLP1* or GLP-1* or glucagon-like peptide* or incretin*):ti,ab,kw (Word variations have been searched) | 7123 | 7583 | 7772 |
| 5 | #1 or #2 or #3 or #4 | 7833 | 10543 | 10809 |
| 6 | MeSH descriptor: [Pneumonia, Aspiration] this term only | 398 | 402 | 402 |
| 7 | MeSH descriptor: [Respiratory Aspiration] explode all trees | 651 | 694 | 715 |
| 8 | (aspirat* or regurgitat*):ti,ab,kw (Word variations have been searched) | 16468 | 17321 | 17625 |
| 9 | MeSH descriptor: [Gastrointestinal Contents] this term only | 163 | 165 | 165 |
| 10 | MeSH descriptor: [Gastric Emptying] this term only | 1812 | 1829 | 1833 |
| 11 | ((gastric or stomach or gastrointestinal) NEAR/2 (content* or volume* or residual or residue or empty*)):ti,ab,kw (Word variations have been searched) | 7228 | 7587 | 7610 |
| 12 | MeSH descriptor: [Ultrasonography] this term only | 6503 | 6592 | 6632 |
| 13 | MeSH descriptor: [Ultrasonic Therapy] this term only | 948 | 963 | 974 |
| 14 | MeSH descriptor: [Endoscopy, Digestive System] explode all trees | 7571 | 7709 | 7768 |
| 15 | MeSH descriptor: [Gastroscopy] this term only | 1079 | 1098 | 1104 |
| 16 | (ultrasound or sonograph* or ultrasonograph* or endoscop* or esophagogastroduodenoscop* or oesophagogastroduodenoscop* or gastroscop*):ti,ab,kw (Word variations have been searched) | 91262 | 96588 | 98436 |
| 17 | #6 or# 7 or #8 or #9 or #10 or #11 or #12 or #13 or #14 or #15 or #16 | 111837 | 118091 | 120196 |
| 18 | #5 AND #17 with Publication Year from 2005 to 2024, in Trials | 792 | 892 | 902 |
| 19 | #18 with Cochrane Library publication date Between Mar 2024 and Oct 2024 | NA | 48 | NA |
| 19 | #18 with Cochrane Library publication date Between Oct 2024 and Jan 2025 | NA | NA | 12 |

| **ClinicalTrials.gov** | | |
| --- | --- | --- |
| **Search Number** | **Search Strategy** | **Initial Search (19 Mar 2024)** |
| 1 | (GLP-1 OR GLP 1 OR glucagon-like peptide OR semaglutide OR tirzepatide OR liraglutide OR exenatide OR dulaglutide) AND (aspiration OR regurgitation OR gastric content OR gastric volume OR gastric residue OR ultrasound OR sonography OR endoscopy) | 19 |

| **ClinicalTrials.gov** | | |
| --- | --- | --- |
| **Search Number** | **Search Strategy** | **Updated Search (21 Oct 2024)** |
| 1 | (GLP-1 OR GLP 1 OR glucagon-like peptide OR semaglutide OR tirzepatide OR liraglutide OR exenatide OR dulaglutide) AND (aspiration OR regurgitation OR gastric content OR gastric volume OR gastric residue OR ultrasound OR sonography OR endoscopy) \| First posted from 03/19/2024 to 10/21/2024 | 6 |

| **ClinicalTrials.gov** | | |
| --- | --- | --- |
| **Search Number** | **Search Strategy** | **Updated Search (13 Jan 2025)** |
| **1** | (GLP-1 OR GLP 1 OR glucagon-like peptide OR semaglutide OR tirzepatide OR liraglutide OR exenatide OR dulaglutide) AND (aspiration OR regurgitation OR gastric content OR gastric volume OR gastric residue OR ultrasound OR sonography OR endoscopy) \| First posted from 10/21/2024 to 01/13/2025 | 8 |

| **World Health Organization (WHO) International Clinical Trials Registry Platform (ICTRP)** | | |
| --- | --- | --- |
| **Search Number** | **Search Strategy** | **Initial Search (19 Mar 2024)** |
| 1 | (GLP-1 OR GLP 1 OR glucagon-like peptide OR semaglutide OR tirzepatide OR liraglutide OR exenatide OR dulaglutide) AND (aspiration OR regurgitation OR gastric content OR gastric volume OR gastric residue OR ultrasound OR sonography OR endoscopy) | 16 |

| **WHO ICTRP** | | |
| --- | --- | --- |
| **Search Number** | **Search Strategy** | **Updated Search (21 Oct 2024)** |
| 1 | (GLP-1 OR GLP 1 OR glucagon-like peptide OR semaglutide OR tirzepatide OR liraglutide OR exenatide OR dulaglutide) AND (aspiration OR regurgitation OR gastric content OR gastric volume OR gastric residue OR ultrasound OR sonography OR endoscopy) \| Date of Registration is between 19/03/2024 to 21/10/2024 | 1 |

| **WHO ICTRP** | | |
| --- | --- | --- |
| **Search Number** | **Search Strategy** | **Updated Search (13 Jan 2025)** |
| 1 | (GLP-1 OR GLP 1 OR glucagon-like peptide OR semaglutide OR tirzepatide OR liraglutide OR exenatide OR dulaglutide) AND (aspiration OR regurgitation OR gastric content OR gastric volume OR gastric residue OR ultrasound OR sonography OR endoscopy) \| Date of Registration is between 21/10/2024 to 13/01/2025 | 2 |

### **Table S2. Outcome definitions for pulmonary aspiration**

| **Study** | **Definition of pulmonary aspiration** | **Measure** |
| --- | --- | --- |
| Alkabbani 2024^3^ | Pulmonary aspiration on the day of or the day after EGD. | Identified from administrative data (MarketScan Commercial Claims and Encounters, and Optum Clinformatics Data Mart) using ICD-10 codes. |
| Amini 2024^4^ | Aspiration pneumonitis within the first 24 to 48 hours post-EGD. | Identified from administrative data (TriNetx) using ICD-10-CM codes. |
| Barlowe 2024^1^ | Aspiration within 14 days of procedure. | Identified from administrative data (Truven Health Analytics MarketScan) using ICD-9/10 codes. |
| Buddhiraju 2024^5^ | Aspiration within 90 days of TKA. | Identified from administrative data (TriNetx) using ICD-10-CM and SNOMED codes. |
| Klonoff 2024^6^ | Aspiration/pneumonitis defined as any diagnosis of pneumonitis because of: (a) aspiration of solids or liquids, (b) pneumonia of bacterial or unspecified origin, or (c) aspiration of fluid within 72 h of surgery | Identified from EMR using ICD-10 codes. |
| Nadeem 2024^7^ | Aspiration. | Identified by review of endoscopy reports. Review method not clearly defined. |
| Peng 2024^8^ | Aspiration pneumonia within 7 days after endoscopy procedure. | Identified from administrative data (TriNetx) using ICD-10 codes. |
| Santos 2024^9^ | Perioperative broncho-aspiration. | Identified from EMR via chart review. Review method not clearly defined. |
| Welk 2024^10^ | Aspiration pneumonia within 14 days of operation. | Identified from administrative data (multiple linked databases) using ICD10 codes. |
| Wu 2024^11^ | Pulmonary aspiration. | Identified from EMR via manual review of all endoscopy and anaesthesia records. |
| Yeo 2024^12^ | Aspiration pneumonia within a month after the procedure. | Identified from administrative data (TriNetx). Coding system not clearly defined. |

Abbreviations: EGD, Esophagogastroduodenoscopy; EMR, electronic medical record; ICD-9, International Classification of Diseases 9th Revision; ICD-10, International Classification of Diseases 10th Revision; ICD-10-CM, International Classification of Disease, Tenth Revision, and Clinical Modification; SNOMED, Systematized nomenclature of medicine; TKA, Total knee arthroplasty.

### **Table S3. Outcome definitions for residual gastric content**

| **Study** | **Definition of residual gastric content** | **Measure** |
| --- | --- | --- |
| Abu-Freha 2024^13^ | Gastric residue was defined as having either:  1) the diagnosis of “poor preparation” on the EGD report, 2) a description of a poor preparation with residual solid food content being found in the stomach within the procedure report, or  3) the need for a repeat EGD due to a poor preparation at the initial procedure. | Identified via endoscopy reports. Review method not clearly defined. |
| Bi 2021^14^ | Retained gastric food during EGD. | Identified from EMR via chart review. Review method not clearly defined. |
| Chapman 2024^15^ | Gastric content retention. | Identified using the Informatics for Integrating Biology & the Bedside database. Review method not clearly defined. Gastric content assessed by a blinded gastroenterologist using the validated POLPREP scale. |
| Elimihele 2024^16^ | Clinically significant delayed gastric emptying defined as patients with EGD findings of retained food, retained partially digested food, phytobezoars, and other findings suggestive of DGE. | Identified via chart review. Review method not clearly defined. |
| Garza 2024^17^ | Gastric residue was defined as the presence of any solids in the stomach not amenable to clearance with suction through the endoscope. | Identified from EMR via manual review of procedure report and recorded images from the procedure. |
| Jirapinyo 2025^18^ | The presence of residual gastric contents during EGD. | Identified by review of endoscopy reports. Review method not clearly defined. |
| Kobori 2023^19^ | Gastric residue defined as having any solids in the stomach in an EGD. | Identified from EMR. Review method not clearly defined. |
| Korlipara 2024^20^ | Retained solid gastric content during EGD, defined as solid contents on photo documentation. | Identified by review of endoscopy and anaesthesia reports. Review method not clearly defined. |
| Nadeem 2024^7^ | Retained gastric contents during EGD. | Identified by review of endoscopy reports. Review method not clearly defined. |
| Nasser 2024^21^ | Retained solid gastric content during EGD when EGD was combined with colonoscopy and in colonoscopy alone. | Identified via endoscopy reports. Review method not clearly defined. |
| Nersessian 2024^22^ | Increased residual gastric content was defined as any solid content or > 1.5 ml.kg-1 of clear fluids. | Gastric ultrasound as described by Perlas and colleagues,^12^ performed by a blinded assessor. |

Abbreviations: DGE, delayed gastric emptying; EGD, esophagogastroduodenoscopy; EMR, electronic medical record; GLP-1 RA, glucagon-like peptide-1 receptor agonist.

**Table S3. Outcome definitions for residual gastric content (continued)**

| **Study** | **Definition of residual gastric content** | **Measure** |
| --- | --- | --- |
| Phan 2024^2^ | Retained gastric contents on EGD. | Identified by review of endoscopy reports. Review method not clearly defined. |
| Pinto 2024^23^ | Residual gastric content at risk of pulmonary aspiration (estimated volume >1.5 mLkg-1 or an ultrasound image suggestive of solid content). | Gastric ultrasound as described by Perlas and colleagues, ^12,15^ performed by an unblinded assessor. |
| Queiroz 2024^24^ | Full stomach defined as solid or fluid content >1.5mL/kg. | Gastric ultrasound as described by Perlas and colleagues, ^12^ performed by a blinded assessor. |
| Robalino Gonzaga 2024^25^ | Gastric food retention, any solid food visualised in the esophagus or stomach during EGD. | Identified via endoscopy reports. Review method not clearly defined. |
| Santos 2024^9^ | Increased residual gastric contents defined as any solid content from the esophagus to the pylorus or  >0.8 mL/Kg of fluid content. | Identified from EMR via chart review. RGC volume reported with predefined categorisation during endoscopy. |
| Sen 2024^26^ | Residual gastric contents defined as solids, thick liquids, or >1.5mL/kg of clear liquids. | Gastric ultrasound as described by Perlas and colleagues, ^12^ performed by a blinded assessor. |
| Sherwin 2023^27^ | Residual gastric contents defined as clear fluid contents >1.5mL/kg or the presence of solids. | Gastric ultrasound as described by Perlas and colleagues, ^12^ performed and reviewed by blinded assessors. |
| Stark 2022^28^ | Food retention during EGD. | Identified from EMR via review of EGD reports. Positive EGD findings validated by two independent investigators. |
| Wu 2024^11^ | Residual gastric contents at the time of EGD. | Identified from EMR via manual review of all endoscopy and anaesthesia records. |

Abbreviations: EGD, esophagogastroduodenoscopy; EMR, electronic medical record; GLP-1 RA, glucagon-like peptide-1 receptor agonist; RGC, residual gastric contents.

### **Table S4. Data requested from study authors**

| **Study** | **Data requested for included studies** | **Data provided** |
| --- | --- | --- |
| Bi 2021^14^ | - Clarification of event rates for patients using GLP-1 RA. - Clarification of covariates. | No |
| Korlipara 2024^20^ | - Clarification of semaglutide type (i.e., daily or weekly formulation). | Yes |
| Phan 2024^2^ | - Clarification of covariates. | No |
| Sen 2024^26^ | - Request for adjusted odds ratio from their sensitivity analysis using a multivariate logistic regression model, which was initially represented as a prevalence ratio. - Request for adjusted odds ratio from a logistic regression model with days since the last dose as a binary variable (i.e., <=7 days since last dose and >7 days since last dose) rather than as a continuous variable. | Yes |
| Welk 2024^10^ | - Clarification of covariates. | Yes |

Abbreviations: GLP-1 RA, glucagon-like peptide-1 receptor agonist

### **Table S5. Studies identified as ongoing or completed without published results at the time of final search**

| **Study Registration** | **Country** | **Study design** | **Data collection** | **Start date** | **End date** | **Ongoing** | **Outcome** | **Outcome measure** | **Assessing time since last dose** | **Assessing effect of an intervention to reduce aspiration or RGC risk** |
| --- | --- | --- | --- | --- | --- | --- | --- | --- | --- | --- |
| NCT05903482 | United States | Observational | Prospective | 6/6/2023 | 31/12/2024 | No | RGC | GUS | No | No |
| NCT06292065 | Belgium | Observational | Prospective | 16/4/2024 | 31/12/2024 | No | RGC; PA | GUS | Yes | No |
| ChiCTR2400081160 | China | Observational | Prospective | 26/2/2024 | 30/12/2024 | No | RGC | GUS | No | No |
| NCT05875636 | United States | Observational | Prospective | 21/8/2023 | 15/9/2024 | No | RGC; PA | EGD | No | No |
| NCT06154486 | Brazil | Observational | Prospective | 19/6/2023 | 22/8/2023 | No | RGC | GUS | Yes | No |
| NCT05854979 | United States | Observational | Prospective | 9/8/2023 | 15/4/2024 | No | RGC; PA | GUS | No | No |
| NCT06038734 | United States | Observational | Prospective | 10/11/2023 | 17/4/2024 | No | RGC | GUS | No | No |
| NCT06263595 | Canada | Observational | Prospective | 31/5/2024 | 31/1/2025 | Yes | RGC | GUS | No | No |
| NCT06003985 | Canada; United States | Observational | Prospective | 29/8/2023 | 1/7/2025 | Yes | RGC | GUS | No | No |
| NCT06533527 | United States | RCT | Prospective | 31/7/2024 | 1/7/2025 | Yes | RGC; PA | EGD; EUS; ERCP | Yes | Yes |
| NCT06581120 | United States | Observational | Prospective | 8/7/2024 | 1/7/2026 | Yes | RGC | GUS | No | No |
| NCT06388213 | Switzerland | Observational | Prospective | 1/6/2024 | 1/5/2026 | Yes | RGC | GUS | Yes | No |
| NCT06500143 | Canada | Observational | Prospective | 1/9/2024 | 1/3/2025 | Yes | RGC | GUS | Yes | No |
| NCT06420739 | Canada | Observational | Prospective | 25/3/2024 | 1/3/2026 | Yes | RGC | GUS | Yes | No |
| NCT06654219 | United States | RCT | Prospective | 1/11/2024 | 1/11/2025 | Yes | RGC | GUS | No | Yes |
| NCT06659159 | Switzerland | Observational | Prospective | 1/11/2024 | 31/12/2026 | Yes | RGC | GUS | No | No |

Abbreviations: EGD, esophagogastroduodenoscopy; ERCP, endoscopic retrograde cholangiopancreatography; EUS, endoscopic ultrasound; GUS, gastric ultrasound; PA, pulmonary aspiration; RCT, Randomised controlled trial; RGC, residual gastric contents

### **Figure S1. Funnel plot for pulmonary aspiration outcome.**

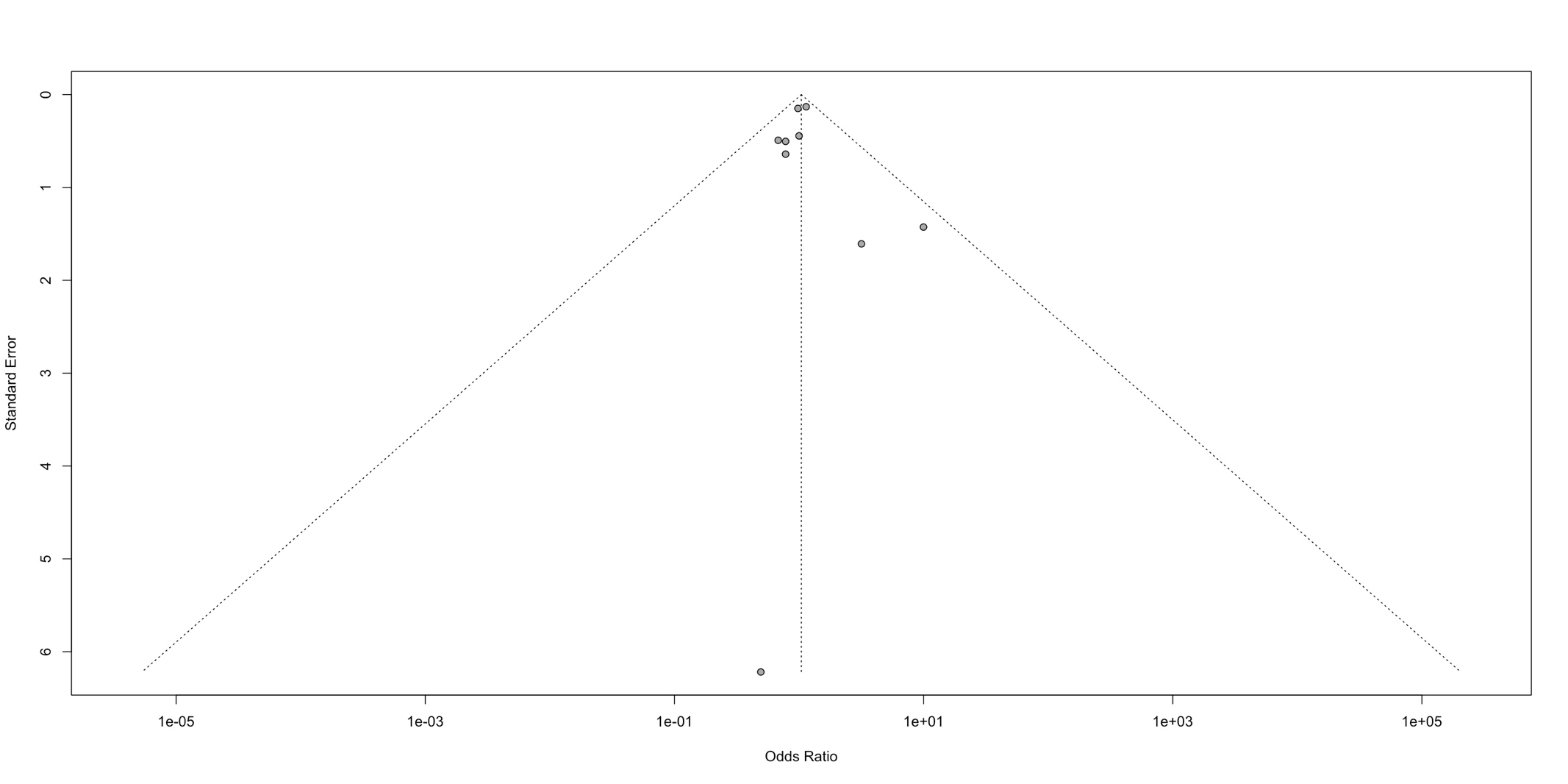

Grey dots indicate point estimate and standard error for each of the included studies assessing pulmonary aspiration.

**Figure S2. Subgroup analysis for pulmonary aspiration outcome comparing studies limited to patients with diabetes to studies not limited to patients with diabetes**

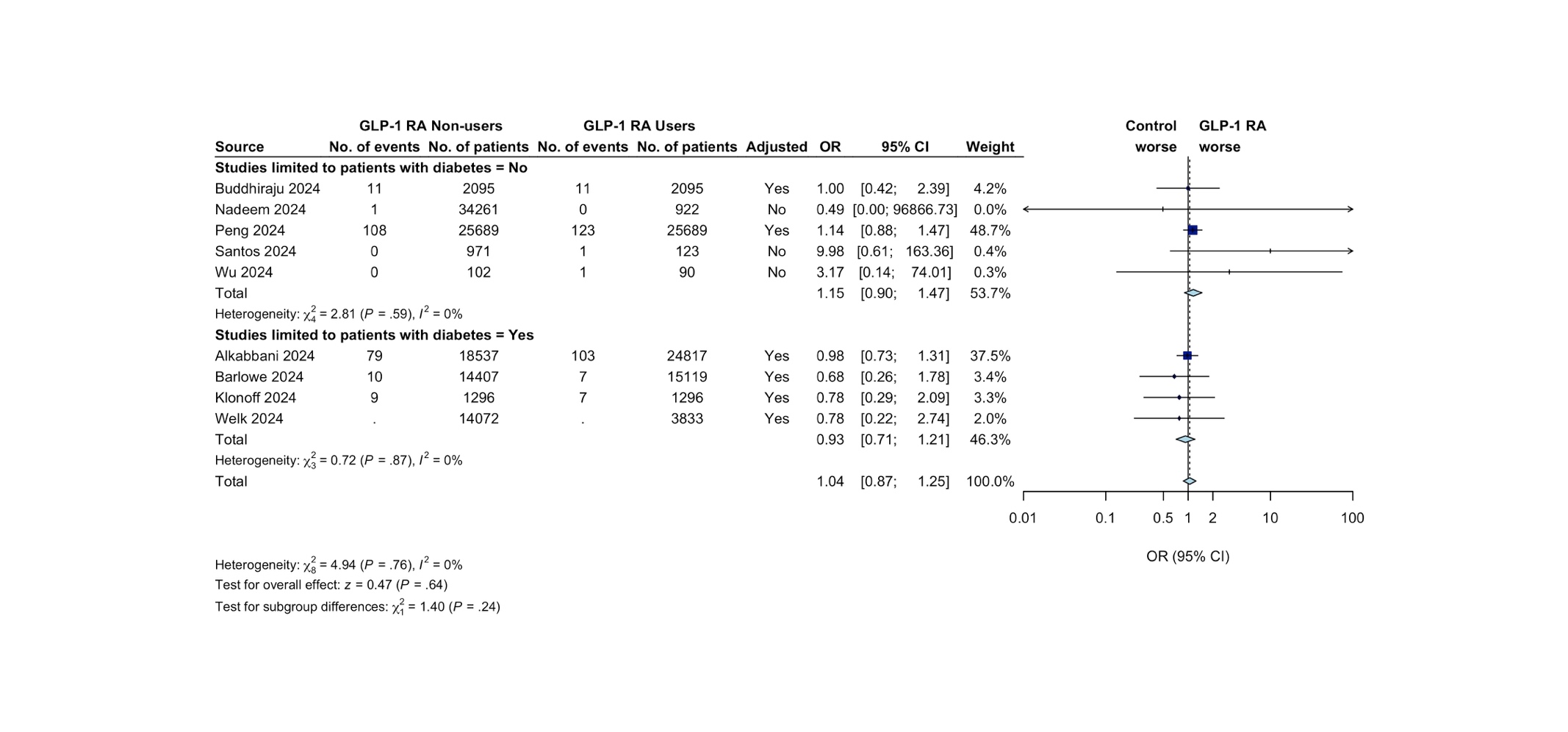

Random-effects model method: restricted maximum likelihood heterogeneity variance estimator. The dark blue boxes represent individual study odds ratio, and the size of the boxes are proportional to study weight in the meta-analysis; the whiskers represent the confidence intervals; light blue diamond represents the overall pooled odds ratio and 95% CI; the dotted vertical line indicates the pooled OR. Abbreviations: GLP-1 RA, Glucagon-Like Peptide-1 Receptor Agonist.

### **Figure S3. Subgroup analysis for pulmonary aspiration outcome comparing studies limited to patients undergoing upper endoscopy procedures to all other studies.**

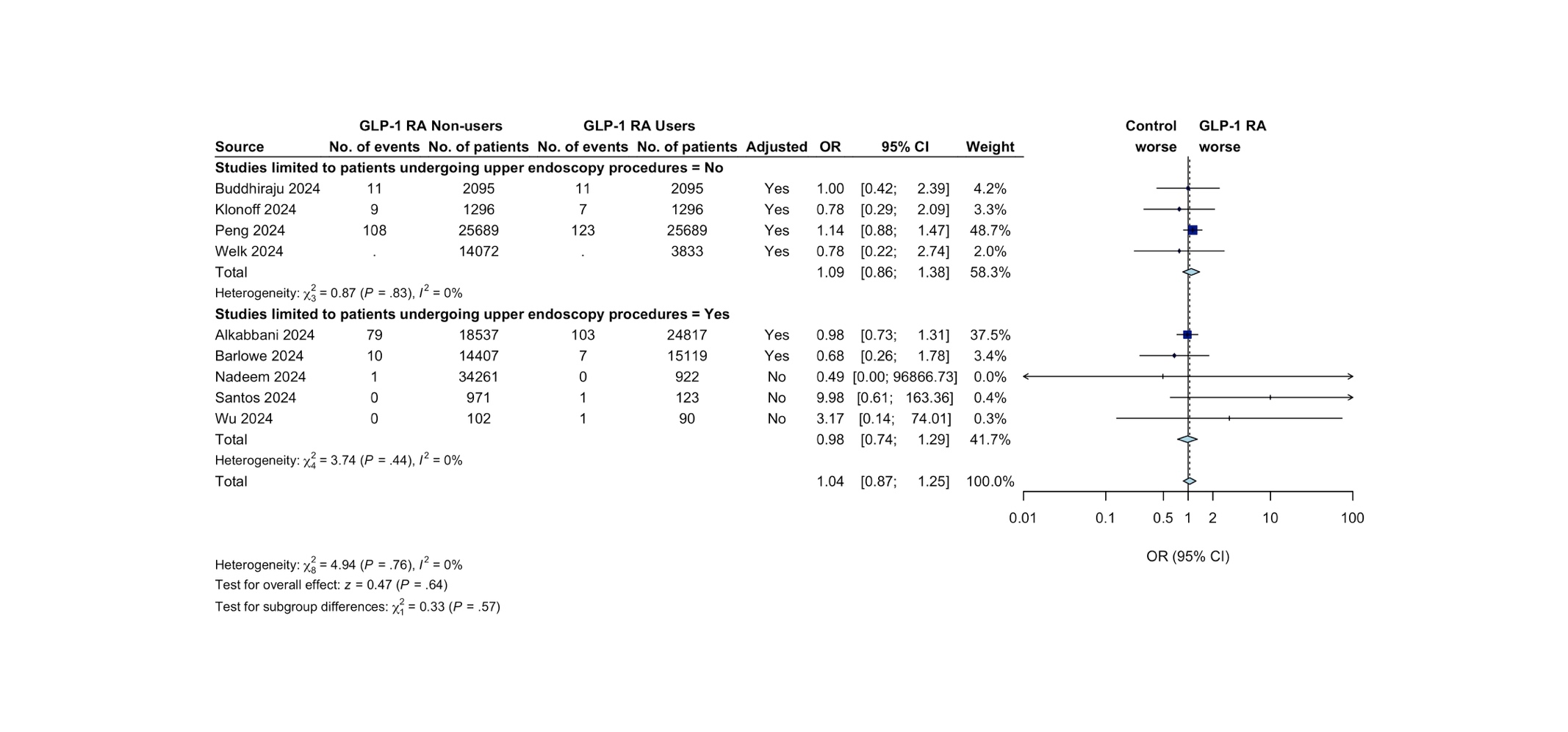

Random-effects model method: restricted maximum likelihood heterogeneity variance estimator. The dark blue boxes represent individual study odds ratio, and the size of the boxes are proportional to study weight in the meta-analysis; the whiskers represent the confidence intervals; light blue diamond represents the overall pooled odds ratio and 95% CI; the dotted vertical line indicates the pooled OR. Abbreviations: GLP-1 RA, Glucagon-Like Peptide-1 Receptor Agonist.

### **Figure S4. Subgroup analysis for pulmonary aspiration outcome comparing studies limited to once-weekly formulations to studies not limited to once-weekly formulations.**

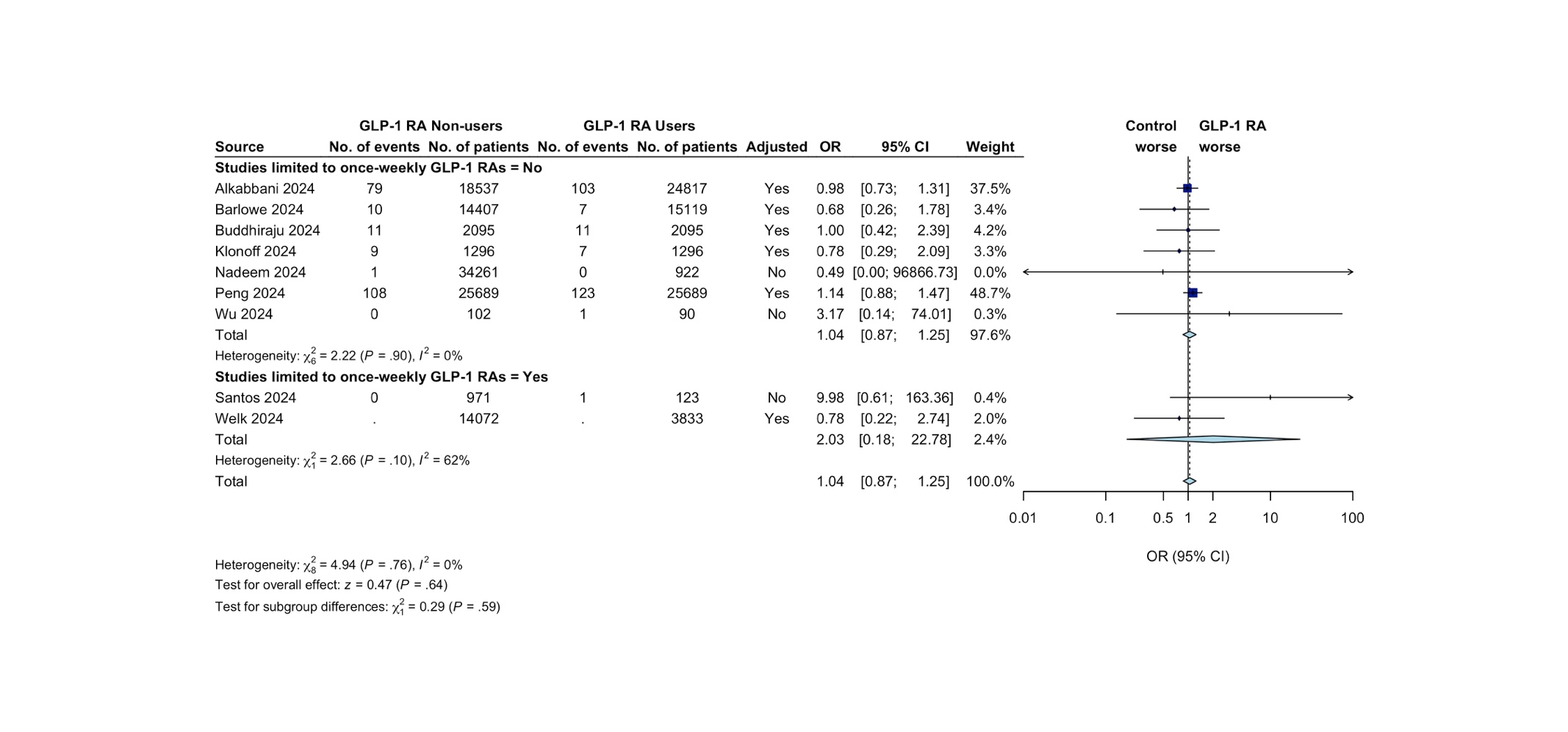

Random-effects model method: restricted maximum likelihood heterogeneity variance estimator. The dark blue boxes represent individual study odds ratio, and the size of the boxes are proportional to study weight in the meta-analysis; the whiskers represent the confidence intervals; light blue diamond represents the overall pooled odds ratio and 95% CI; the dotted vertical line indicates the pooled OR. Abbreviations: GLP-1 RA, Glucagon-Like Peptide-1 Receptor Agonist.

### **Table S6. Modified Quality in Prognosis Studies (QUIPS) tool assessment of pulmonary aspiration**

| **Study** | **Study Participation** | **Study Attrition** | **Prognostic factor measurement** | **Outcome measurement** | **Study confounding** | **Statistical analysis and reporting** | **Overall Bias*** |
| --- | --- | --- | --- | --- | --- | --- | --- |
| Alkabbani 2024^3^ | Moderate | Low | Moderate | Low | Low | Low | Moderate |
| Amini 2024^4^ | Moderate | Moderate | Moderate | Low | Moderate | Moderate | Moderate |
| Barlowe 2024^1^ | Moderate | Moderate | Moderate | Low | Moderate | Moderate | Moderate |
| Buddhiraju 2024^5^ | Moderate | Moderate | Moderate | Moderate | Moderate | Moderate | Moderate |
| Klonoff 2024^6^ | Moderate | Moderate | Moderate | Low | Moderate | Moderate | Moderate |
| Nadeem 2024^7^ | Moderate | Moderate | Moderate | Moderate | High | Moderate | High |
| Peng 2024^8^ | Moderate | Moderate | Moderate | Low | Low | Moderate | Moderate |
| Santos 2024^9^ | Moderate | Moderate | Low | Moderate | High | Moderate | High |
| Welk 2024^10^ | Moderate | Moderate | Low | Low | Low | Moderate | Moderate |
| Wu 2024^11^ | Moderate | Moderate | Low | Moderate | High | Moderate | High |
| Yeo 2024^12^ | Moderate | Moderate | Moderate | Moderate | Moderate | Moderate | Moderate |

Green indicates low risk of bias; Yellow indicates moderate risk of bias; Red indicates high risk of bias.

* Studies were rated as low risk of bias if all or most of the 6 domains were marked as low; Studies were rated as moderate risk of bias if most of the 6 domains were marked as moderate and none were marked as high; Studies were rated as high risk of bias if at least one domain was marked as high.

### **Table S7. Adjusted variables in included studies assessing pulmonary aspiration**

| **Study** | **Age** | **Sex** | **BMI** | **Diabetes*** | **ASA-PS** | **Medication** | **Other comorbidities** | **Other variables** |
| --- | --- | --- | --- | --- | --- | --- | --- | --- |
| Alkabbani 2024^3^ | Yes | Yes | Yes | NA | No | Medication use; Statins; Opioids; Metformin; Insulins; 2nd generation Sulfonylureas; Thiazolidinediones; DPP-4 inhibitors; Antiemetics; Antidepressants; Antiparkinsonian medications; Lithium; Antipsychotics; Beta blockers; CCB; Thiazide diuretics; Loop diuretics; MRA; Gabapentinoids; Benzodiazepines; H2 blocker; PPI. | Diabetic neuropathy, Diabetic retinopathy, CKD; Hyperlipidaemia; Hypertension; Coronary artery disease; Hemorrhagic stroke; Ischemic stroke; Unstable angina; Stable angina; Acute myocardial Infarction; Heart failure; Peripheral artery disease; NAFL and NASH; Obstructive sleep apnoea; Osteoarthritis; GERD; GI tract cancer; Peptic ulcer disease; Inflammatory bowel disease; Gastritis and duodenitis; Esophagitis; Helicobacter Pylori; GI Bleeding; Metaplasia; Gastroparesis; Other diseases of the esophagus; Other gastroduodenal diseases. | Length of stay in hospital; Number of endocrinologist visits; Number of hospitalizations; Number of distinct medications; Race; Region; Frailty index; Smoking; Alcohol abuse or dependence; Drug abuse or dependence. |
| Amini 2024^4^ | Yes | Yes | No | Yes | No | None | Obesity | Nicotine dependence |
| Barlowe 2024^1^ | Yes | Yes | No | NA | No | No | History of COPD; Asthma; Gastroparesis | CCI score |

Abbreviations: ASA-PS, American Society of Anesthesiologists Physical Status; BMI, Body mass index; CCB, Calcium channel blockers; CCI, Charlson comorbidity index; CKD, Chronic kidney disease; COPD, Chronic obstructive pulmonary disease; DPP-4, Dipeptidyl peptidase 4; GERD, Gastroesophageal reflux disease; GI, gastrointestinal; H2 blocker, Histamine type-2 receptor antagonists; MRA, Mineralocorticoid receptor antagonists; NA, Not applicable; NAFL, Non-alcoholic fatty liver disease; NASH, Non-alcoholic steatohepatitis; PPI, Proton pump inhibitor.

* Studies marked as NA restricted their inclusion criteria to patients with diabetes.

**Table S7. Adjusted variables in included studies assessing pulmonary aspiration (continued)**

| **Study** | **Age** | **Sex** | **BMI** | **Diabetes*** | **ASA-PS** | **Medication** | **Other comorbidities** | **Other variables** |
| --- | --- | --- | --- | --- | --- | --- | --- | --- |
| Buddhiraju 2024^5^ | Yes | Yes | No | Yes | No | None | Obesity; Hypertension; Ischemic heart disease; Acute kidney failure; CKD; Mood (affective) disorders such as depression, and anxiety, dissociative, stress-related, somatoform, and other nonpsychotic mental disorders. | Race; Ethnicity, HbA1C. |
| Klonoff 2024^6^ | Yes | Yes | No | NA | No | Medication codes | Not specified | CCI score; Treatment year; Diagnostic codes; Procedure codes; Number and frequency of encounters in the EHR database |
| Nadeem 2024^7^ | No | No | No | No | No | None | None | None |
| Peng 2024^8^ | Yes | Yes | Yes | Yes | No | Glucose-lowering and gastrointestinal agents. | Morbid obesity; GERD; Gastroparesis; Dysphagia; Hiatal hernia; Achalasia; Dyskinesia of esophagus; Oesophageal diverticulum; Acquired absence of part of stomach; Parkinson disease; Cerebral infarction; Multiple sclerosis. | Race; HbA1C. |
| Santos 2024^9^ | No | No | No | No | No | None | None | None |

Abbreviations: ASA-PS, American Society of Anesthesiologists physical status; BMI, Body mass index; CCI, Charlson comorbidity index; CKD, Chronic kidney disease; EHR, Electronic health record; GERD, Gastroesophageal reflux disease; GLP-1 RA, Glucagon-like peptide-1 receptor agonist; HbA1C, Haemoglobin A1C.

* Studies marked as NA restricted their inclusion criteria to patients with diabetes.

**Table S7. Adjusted variables in included studies assessing pulmonary aspiration (continued)**

| **Study** | **Age** | **Sex** | **BMI** | **Diabetes*** | **ASA-PS** | **Medication** | **Other comorbidities** | **Other variables** |
| --- | --- | --- | --- | --- | --- | --- | --- | --- |
| Welk 2024^10^ | Yes | Yes | No | NA | No | PPI; Narcotic; Insulin. | Conditions present in prior 5 years (All-cause pneumonia; Aspiration pneumonia; Morbid obesity (BMI >45); Stroke; Dementia; Congestive heart failure; COPD; Hypertension; Coronary artery disease with angina; Atrial fibrillation/flutter; Seizure; Alcoholism; Bariatric surgery). | Long-term care resident; CCI score; Duration of diabetes; Surgical procedure; Post-operative length of stay; Surgery in 2020; Surgery in 2021; Surgery in 2022; Surgery in 2023; Diabetes management visit; Family practise diabetes management visit. |
| Wu 2024^11^ | No | No | No | No | No | None | None | None |
| Yeo 2024^12^ | Yes | Yes | Yes | Yes | No | Opioid analgesics; Metformin; Ondansetron; Insulin; Fentanyl; Midazolam; Propofol; Diphenhydramine; Promethazine; Opioid antagonists; Glipizide; Lorazepam; Metoclopramide; Sitagliptin; TCA; Diazepam; Zolpidem; Empagliflozin; Prochlorperazine; Alprazolam; Glimepiride; Meclizine; Scopolamine; Pioglitazone; Canagliflozin; Dapagliflozin; Glyburide; Linagliptin; Olanzapine; Dimenhydrinate; Droperidol; Saxagliptin; Alogliptin; Palonosetron; Fosaprepitant; Chlorpromazine; Chlorpromazine; Acarbose; Granisetron; Dolasetron. | Overweight/obese; CKD; Cerebral infarction; Other chronic pancreatitis; Dementia; Vascular dementia; Alzheimer disease; Gastroparesis; Dysphagia. | Race; HbA1C; Fasting glucose. |

Abbreviations: ASA-PS, American Society of Anesthesiologists physical status; BMI, Body mass index; CCI, Charlson comorbidity index; COPD, Chronic obstructive pulmonary disease; CKD, Chronic kidney disease; GLP-1 RA, glucagon-like peptide-1 receptor agonist; HbA1C, Haemoglobin A1C; PPI, Proton pump inhibitor; TCA, Tricyclic antidepressants.

* Studies marked as NA restricted their inclusion criteria to patients with diabetes

### **Figure S5. Funnel plot for residual gastric contents outcome.**

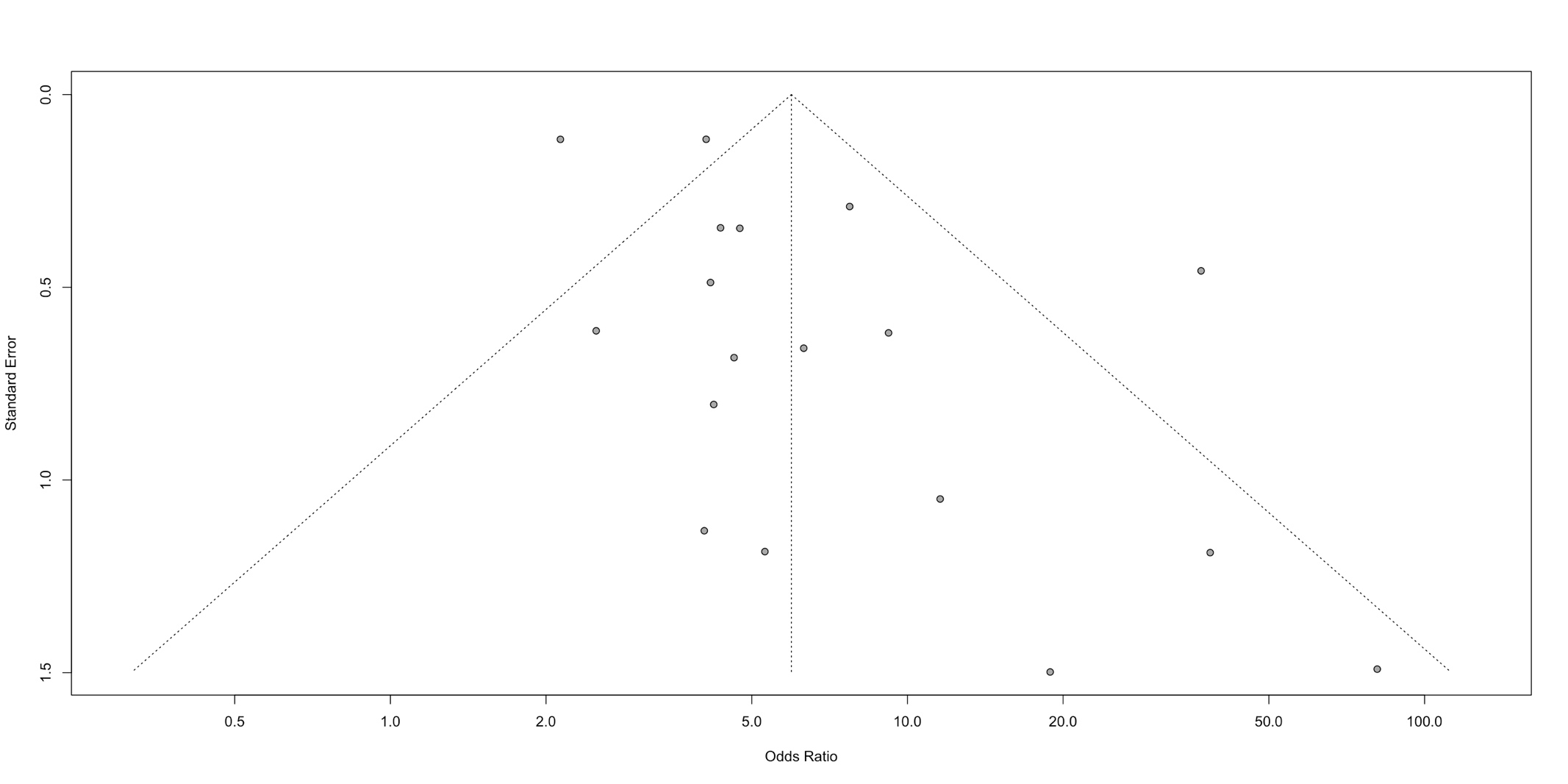

Grey dots indicate point estimate and standard error for each of the included studies assessing pulmonary aspiration.

### **Figure S6. Significance plot for residual gastric contents outcome.**

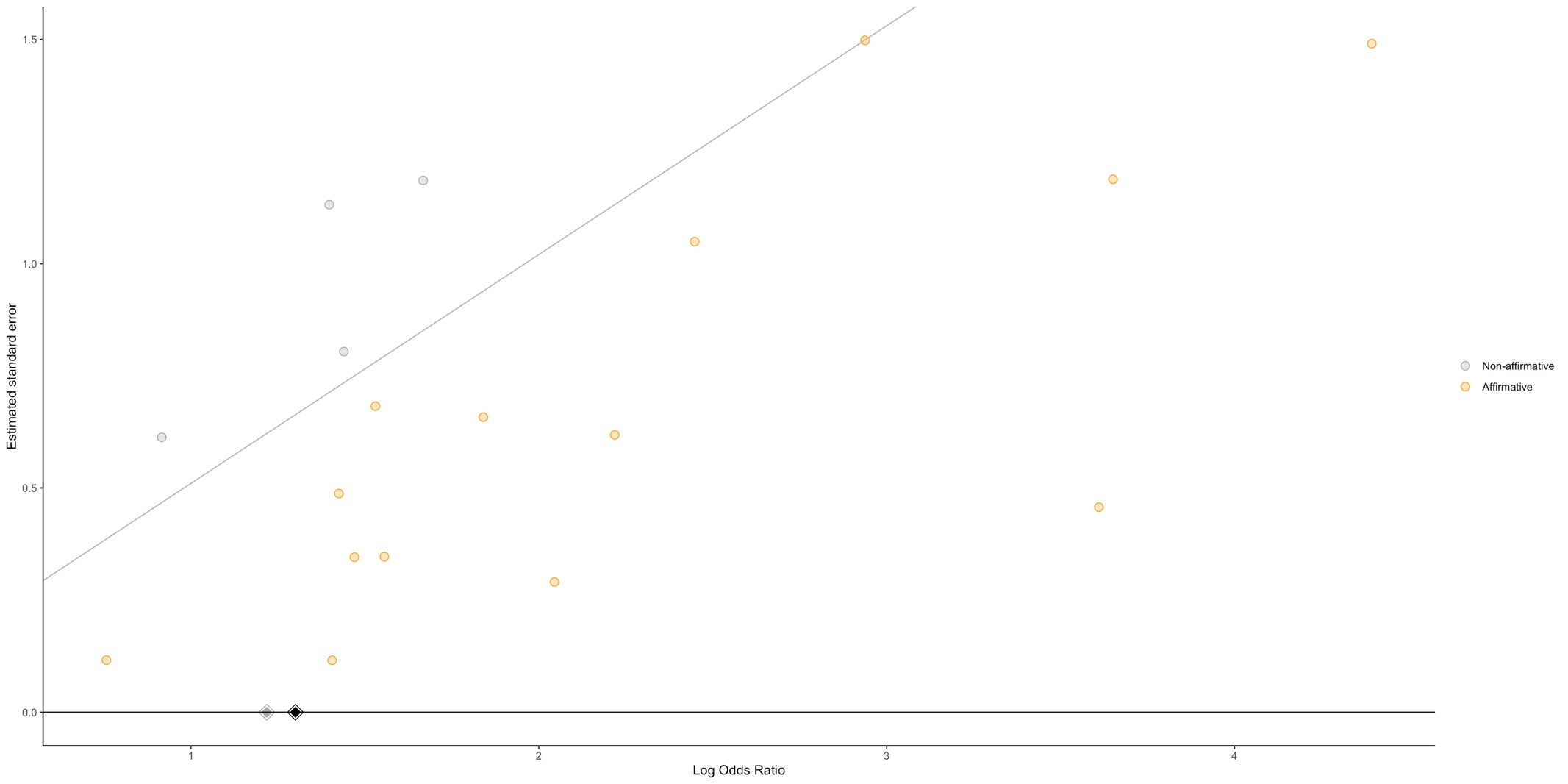

Grey dot indicates non-affirmative studies; orange dot indicates affirmative studies; black diamond indicates estimate from meta-analysis for all studies; grey diamond indicates sensitivity analysis of non-affirmative studies.

### **Figure S7. Sensitivity analysis of non-affirmative studies for residual gastric contents outcome.**

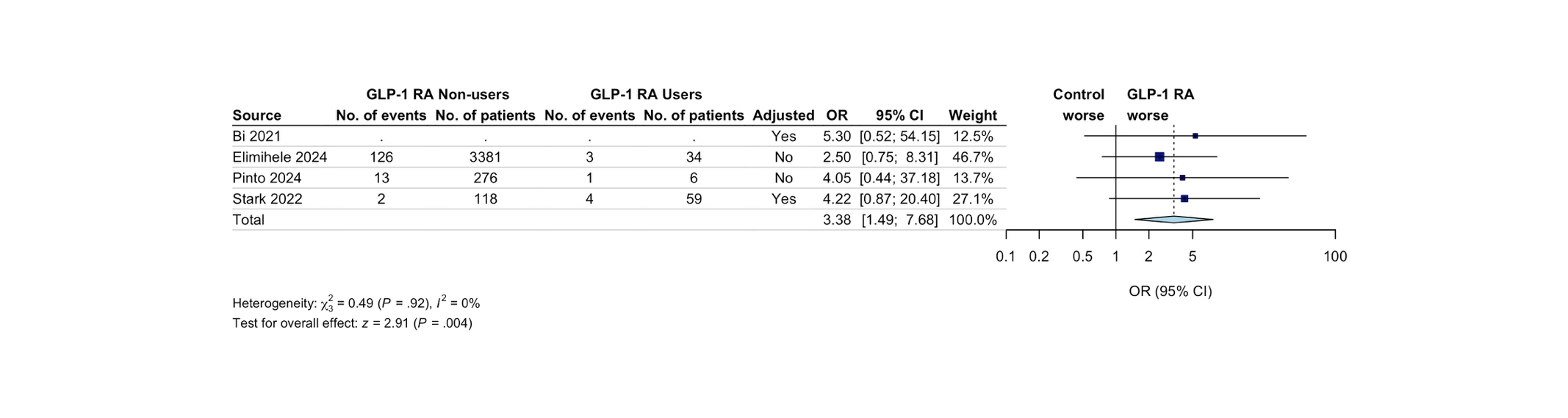

Random-effects model with restricted maximum likelihood heterogeneity variance estimator. The dark blue boxes represent individual study odds ratio, and the size of the boxes are proportional to study weight in the meta-analysis; the whiskers represent the confidence intervals; light blue diamond represents the overall pooled odds ratio and 95% CI; the dotted vertical line indicates the pooled OR. Adjusted indicates adjusted ORs. Abbreviations: GLP-1 RA, Glucagon-Like Peptide-1 Receptor Agonist.

**Figure S8. Subgroup analysis for residual gastric contents outcome comparing studies limited to patients undergoing upper endoscopy procedures to all other studies**
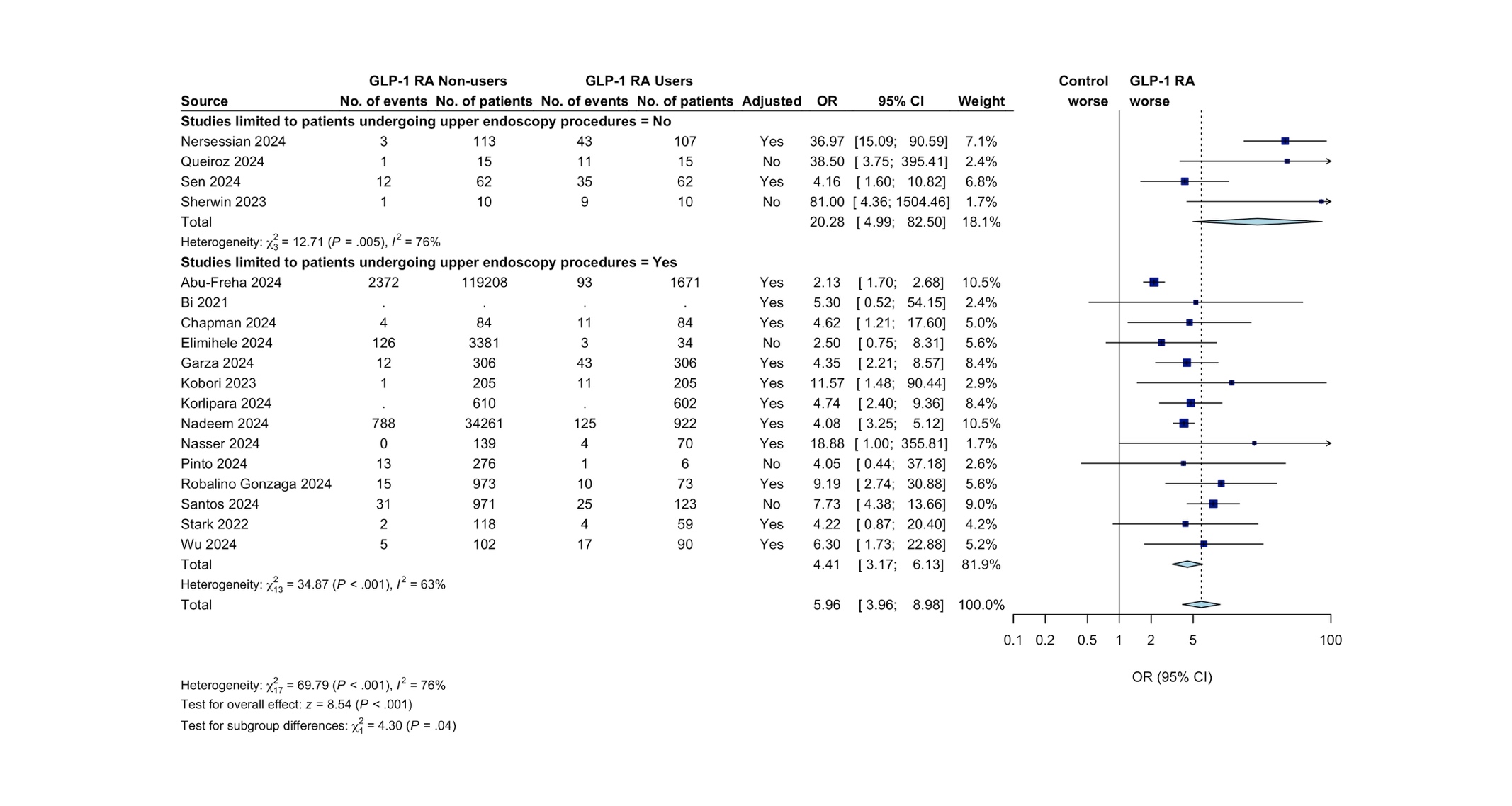

Random-effects model method: restricted maximum likelihood heterogeneity variance estimator. The dark blue boxes represent individual study odds ratio, and the size of the boxes are proportional to study weight in the meta-analysis; the whiskers represent the confidence intervals; light blue diamond represents the overall pooled odds ratio and 95% CI; the dotted vertical line indicates the pooled OR. Abbreviations: GLP-1 RA, Glucagon-Like Peptide-1 Receptor Agonist.

### **Figure S9. Subgroup analysis for residual gastric contents outcome comparing studies limited to patients with diabetes to studies not limited to patients with diabetes**

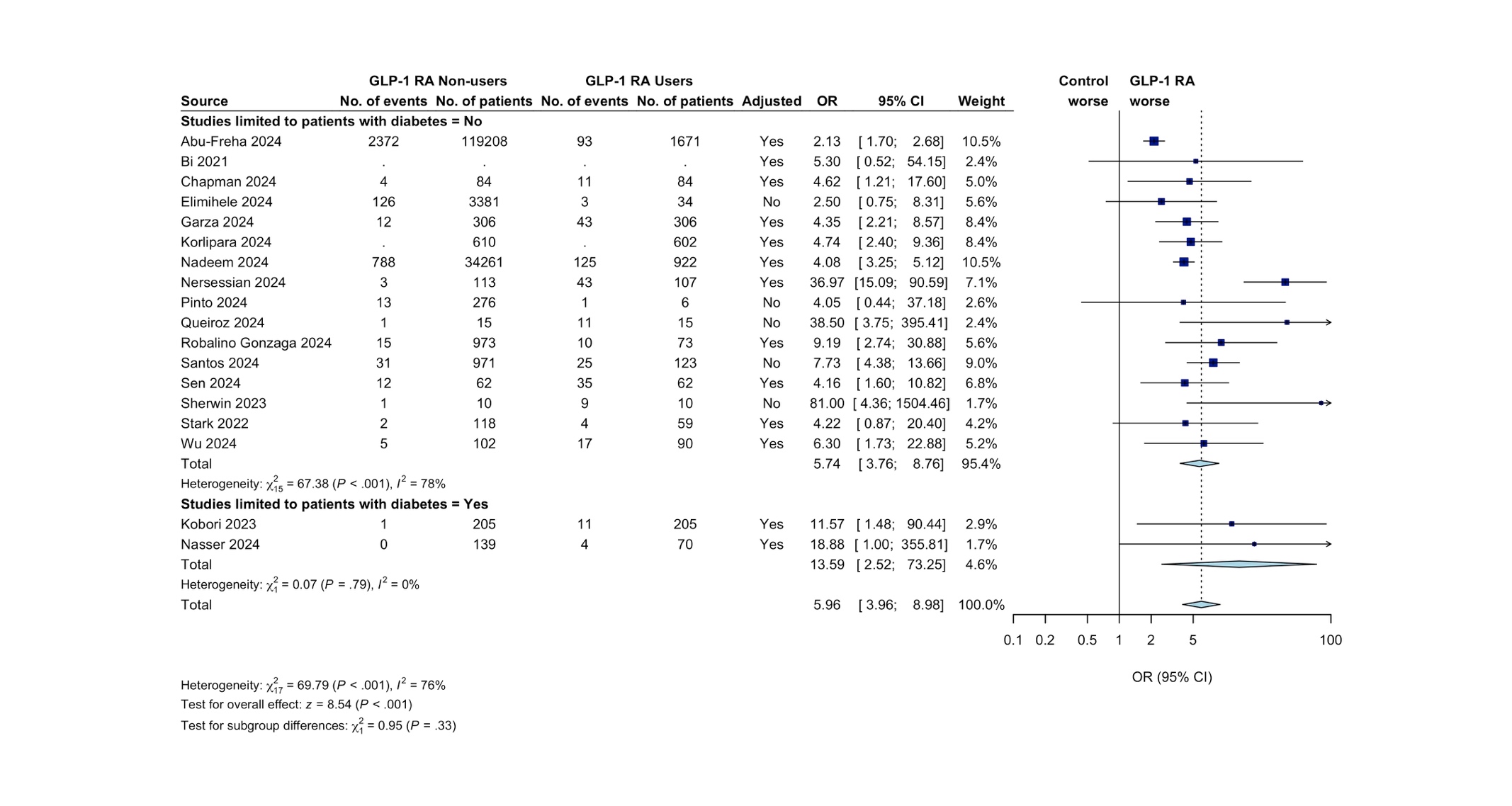

Random-effects model method: restricted maximum likelihood heterogeneity variance estimator. The dark blue boxes represent individual study odds ratio, and the size of the boxes are proportional to study weight in the meta-analysis; the whiskers represent the confidence intervals; light blue diamond represents the overall pooled odds ratio and 95% CI; the dotted vertical line indicates the pooled OR. Abbreviations: GLP-1 RA, Glucagon-Like Peptide-1 Receptor Agonist

### **Figure S10. Subgroup analysis for residual gastric contents outcome comparing studies limited to once-weekly formulations to studies not limited to once-weekly formulations.**

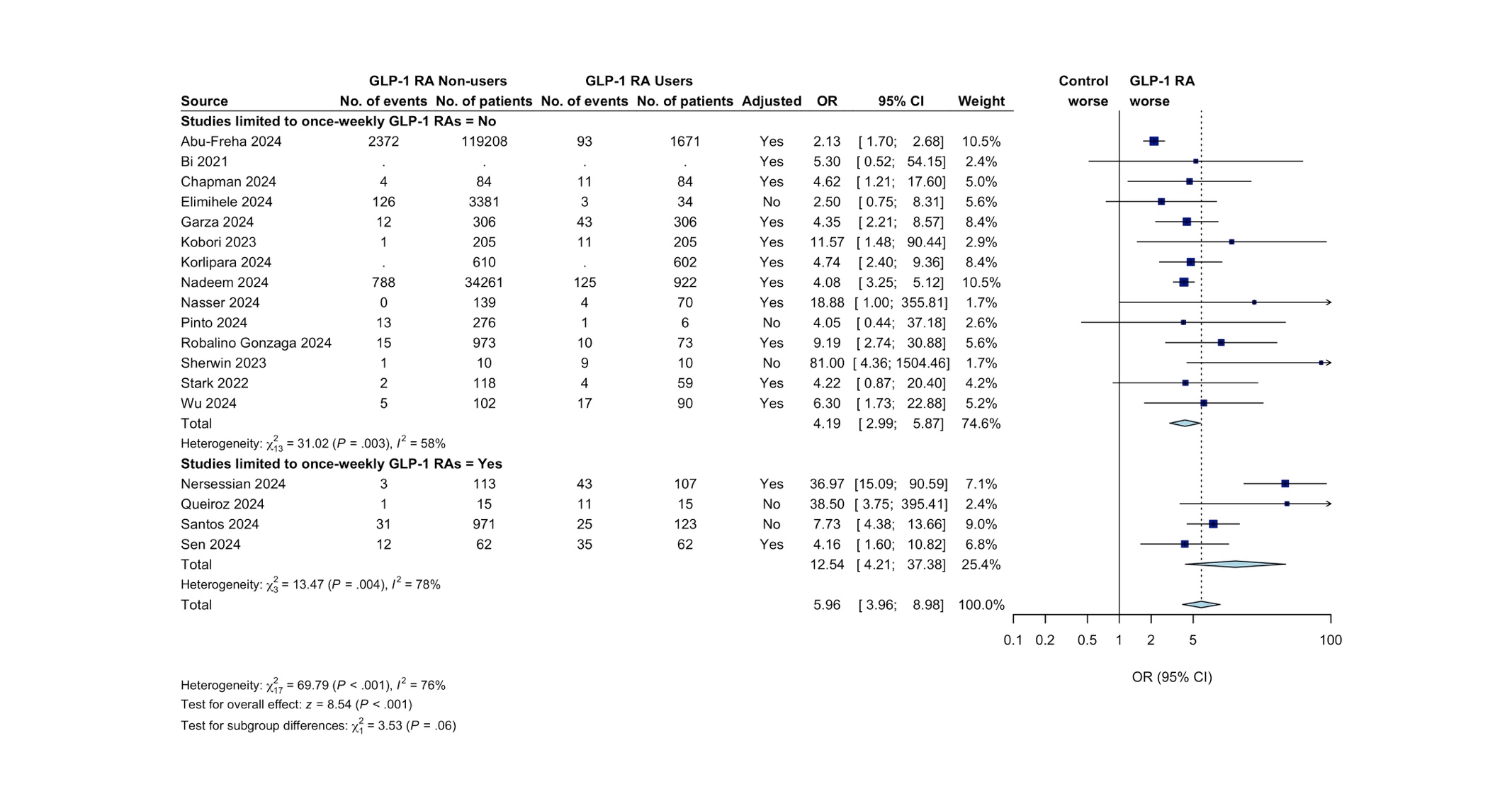

Random-effects model method: restricted maximum likelihood heterogeneity variance estimator. The dark blue boxes represent individual study odds ratio, and the size of the boxes are proportional to study weight in the meta-analysis; the whiskers represent the confidence intervals; light blue diamond represents the overall pooled odds ratio and 95% CI; the dotted vertical line indicates the pooled OR. Abbreviations: GLP-1 RA, Glucagon-Like Peptide-1 Receptor Agonist.

### **Table S8. Modified Quality in Prognosis Studies (QUIPS) tool assessment of residual gastric contents**

| **Study** | **Study Participation** | **Study Attrition^*^** | **Prognostic factor measurement** | **Outcome measurement** | **Study confounding** | **Statistical analysis and reporting** | **Overall Bias**† |
| --- | --- | --- | --- | --- | --- | --- | --- |
| Abu-Freha 2024^13^ | Moderate | NA | Moderate | High | Moderate | Moderate | High |
| Bi 2021^14^ | High | NA | Moderate | High | High | High | High |
| Chapman 2024^15^ | Moderate | NA | Moderate | Low | Moderate | moderate | Moderate |
| Elimihele 2024^16^ | Moderate | NA | Moderate | High | High | High | High |
| Garza 2024^17^ | Moderate | NA | Low | Moderate | Moderate | Moderate | Moderate |
| Kobori 2023^19^ | Moderate | NA | Moderate | High | Moderate | Moderate | High |
| Korlipara 2024^20^ | Moderate | NA | Moderate | High | Moderate | moderate | High |
| Nadeem 2024^7^ | Moderate | NA | Moderate | High | Moderate | Moderate | High |
| Nasser 2024^21^ | Moderate | NA | Moderate | High | Moderate | Moderate | High |
| Nersessian 2024^22^ | High | NA | Moderate | Low | Moderate | Moderate | High |
| Pinto 2024^23^ | Moderate | NA | Moderate | Moderate | High | Low | High |
| Queiroz 2024^24^ | High | NA | Moderate | Low | High | Moderate | High |
| Robalino Gonzaga 2024^25^ | Moderate | NA | Moderate | High | Moderate | Moderate | High |
| Santos 2024^9^ | Moderate | NA | Low | Moderate | High | Moderate | High |
| Sen 2024^26^ | High | NA | Moderate | Low | Low | Low | High |
| Sherwin 2023^27^ | High | NA | Moderate | Low | High | High | High |
| Stark 2022^28^ | Moderate | NA | Moderate | Moderate | High | Moderate | High |
| Wu 2024^11^ | Moderate | NA | Low | Moderate | Moderate | Low | Moderate |

Green indicates low risk of bias; Yellow indicates moderate risk of bias; Red indicates high risk of bias. Abbreviations: NA, not applicable.

* Studies marked as NA due to acute presentation with residual gastric contents, therefore assessment of study attrition is not relevant.

† Studies were rated as low risk of bias if all or most of the 6 domains were marked as low; Studies were rated as moderate risk of bias if most of the 6 domains were marked as moderate and none were marked as high; Studies were rated as high risk of bias if at least one domain was marked as high.

### **Table S9. Adjusted variables in included studies assessing residual gastric contents**

| **Study** | **Age** | **Sex** | **BMI** | **Diabetes*** | **ASA-PS** | **Medication** | **Other comorbidities** | **Other variables** |
| --- | --- | --- | --- | --- | --- | --- | --- | --- |
| Abu-Freha 2024^13^ | Yes | Yes | No | Yes | No | None | None | None |
| Bi 2021^14^ ^b^ | NR | NR | NR | NR | NR | NR | NR | NR |
| Chapman 2024^15^ ^c^ | Yes | Yes | NR | Yes | NR | No | NR | Race |
| Elimihele 2024^16^ | No | No | No | No | No | None | None | None |
| Garza 2024^17^ | No | Yes | Yes | Yes | No | Gastric modulators; Insulin therapy | None | Concomitant colonoscopy; Time of endoscopy; GLP1-RA usage; Diabetes complications. |
| Jirapinyo 2025^18^ | No | No | No | No | No | None | None | None |
| Kobori 2023^19^ | Yes | Yes | No | NA | No | Insulin treatment | None | HbA1c; Fasting blood glucose level |
| Korlipara 2024^20^ ^b^ | NR | NR | NR | NR | NR | NR | NR | NR |
| Nadeem 2024^7^ | Yes | Yes | Yes | Yes | No | None | None | Race/ethnicity |
| Nasser 2024^21^ | Yes | Yes | Yes | No | No | None | None | Procedure |
| Nersessian 2024^22^ | Yes | Yes | Yes | NA | Yes | None | None | None |
| Phan 2024^2^ | NR | NR | NR | NR | NR | NR | NR | NR |
| Pinto 2024^23^ | No | No | No | No | No | None | None | None |
| Queiroz 2024^24^ | No | No | No | No | No | None | None | None |
| Robalino Gonzaga 2024^25^ | Yes | No | No | Yes | No | Antimotility drugs | CKD | Race; Same-day colonoscopy. |
| Santos 2024^9^ | No | No | No | No | No | None | None | None |
| Sen 2024^26^ | Yes | Yes | Yes | Yes | Yes | Opioid use | None | Pain score; Time since last oral intake |
| Sherwin 2023^27^ | No | No | No | No | No | None | None | None |
| Stark 2022^28^ | No | No | No | Yes | No | No | Cirrhosis | None |
| Wu 2024 ^11^ | Yes | Yes | Yes | Yes | Yes | Insulin; Oral diabetic agents | None | Incidence of gastroesophageal reflux symptoms; EGD indication group. |

Abbreviations: ASA-PS, American Society of Anesthesiologists physical status; BMI, Body mass index; CKD, Chronic kidney disease; EGD, Esophagogastroduodenoscopy; GLP-1 RA, Glucagon-like peptide-1 receptor agonist; HbA1C, Haemoglobin A1C; NA, Not applicable; NR, Not reported; PPI, Proton pump inhibitor.

* Studies marked as NA restricted their inclusion criteria to patients with diabetes.

† Variables adjusted for in the multivariate logistic regression were not clearly reported.

‡ Propensity score matching was performed using the variables listed in the table. Variables adjusted for in the multivariate logistic regression were not clearly reported.

### **Table S10. Studies assessing withholding at least one GLP-1 RA dose prior to a procedure on residual gastric contents**

| **Study** | **Exposure** | **No. events** | **No. patients held*** | **No. events** | **No. patients not held**† | **Odds ratio (95% CI)** ‡ | **Adjusted** |
| --- | --- | --- | --- | --- | --- | --- | --- |
| Jirapinyo 2025^18^ | Any GLP-1 RA (daily or weekly-dosed) | 4 | 146 | 31 | 483 | 0.41 (0.14; 1.18) § | No |
| Nersessian 2024^22^ | Once-weekly semaglutide | 7 | 20 | 36 | 87 | 0.76 (0.26–2.06) | No |
| Phan 2024^2^ | Any GLP-1 RA (daily or weekly-dosed) | 18 | 406 | 52 | 409 | 2.86 (1.59 to 5.18) | Yes |
| Santos 2024^9^ | Once-weekly semaglutide | 17 | 91 | 8 | 32 | 0.69 (0.26; 1.80) § | No |
| Sen 2024^26^ | Once-weekly GLP-1 RA | 5 | 7 | 30 | 55 | 2.15 (0.29 to 15.79) | Yes |

Abbreviations: CI, confidence interval; GLP-1 RA, glucagon-like peptide-1 receptor agonist; NR, not reported; OR, odds ratio; PR, prevalence ratio.

* Indicates patients who had their last GLP-1 RA dose either > 7 days (weekly doses) or >1 day (daily doses) prior to measurement of residual gastric contents.

† Indicates patients who had their last GLP-1 RA dose ≤ 7 days (weekly doses) or ≤ 1 day (single doses) prior to measurement of residual gastric contents.

‡	Patients who did not hold their GLP-1 RA prior to procedure were meta-analysed as the control group. Where appropriate, we took the inverse of published estimates to ensure consistency in the direction of the pooled effect estimates.

§ Study did not report effect estimates. Unadjusted odds ratio was calculated using the unadjusted event rates reported in the manuscript.

### **Figure S11. Funnel plot for studies assessing withholding at least one GLP-1 RA dose prior to a procedure on residual gastric contents.**

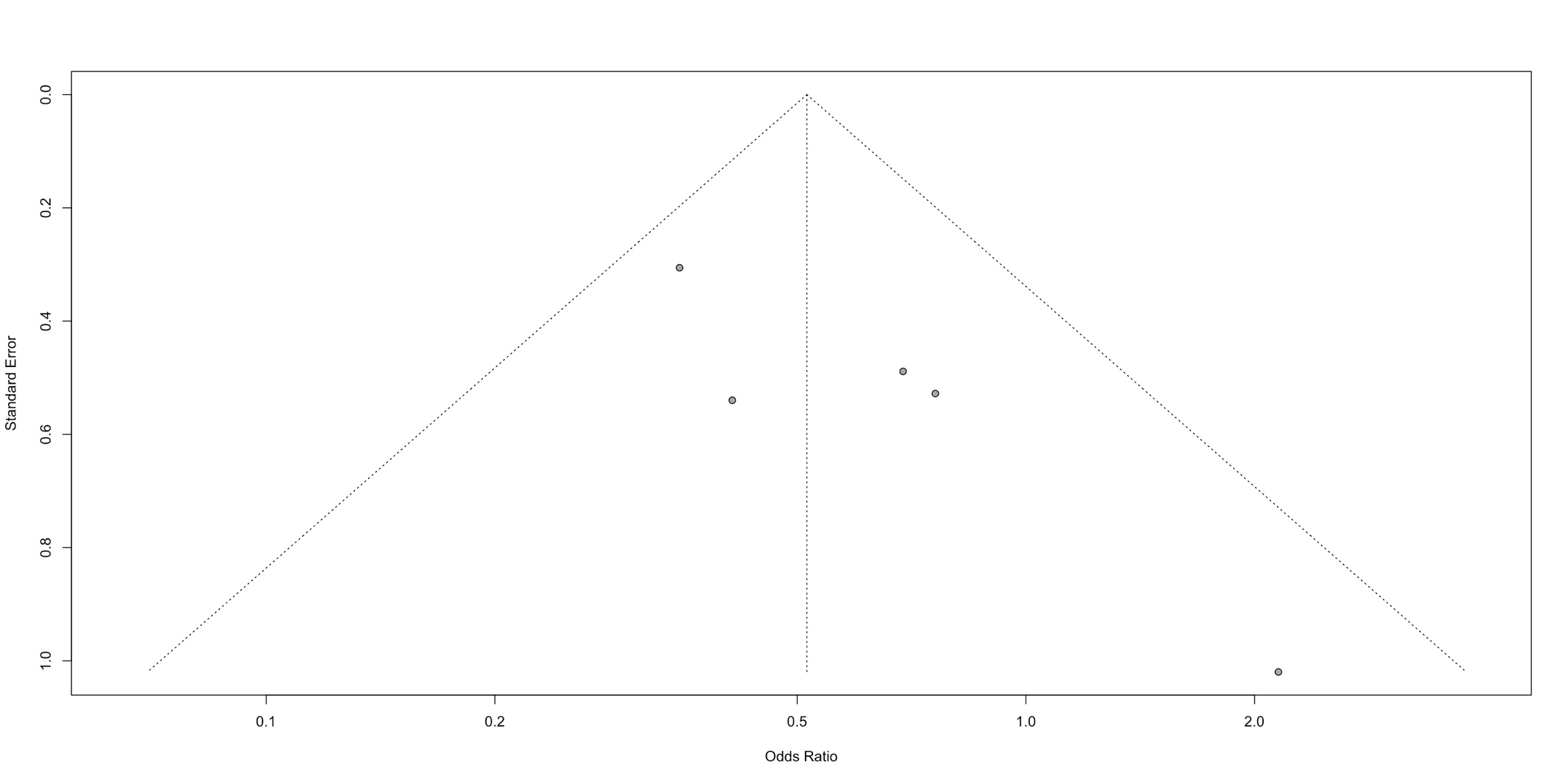

Grey dots indicate point estimate and standard error for each of the included studies assessing pulmonary aspiration.

### **Figure S12. Significance plot for studies assessing withholding at least one GLP-1 RA dose prior to a procedure on residual gastric contents.**

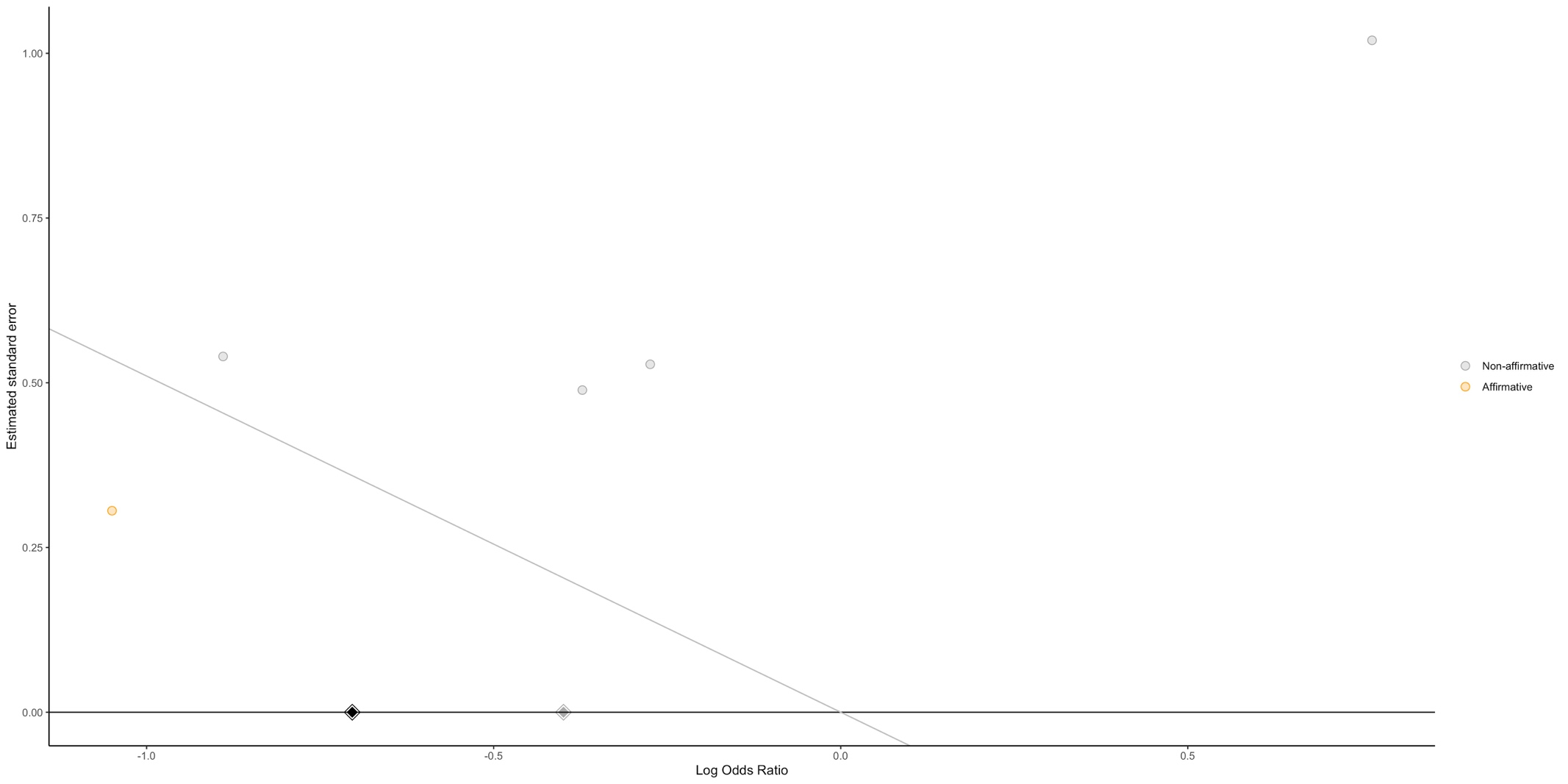

Grey dot indicates non-affirmative studies; orange dot indicates affirmative studies; black diamond indicates estimate from meta-analysis for all studies; grey diamond indicates sensitivity analysis of non-affirmative studies.

### **Figure S13. Sensitivity analysis of non-affirmative studies for studies assessing withholding at least one GLP-1 RA dose prior to a procedure on residual gastric contents.**

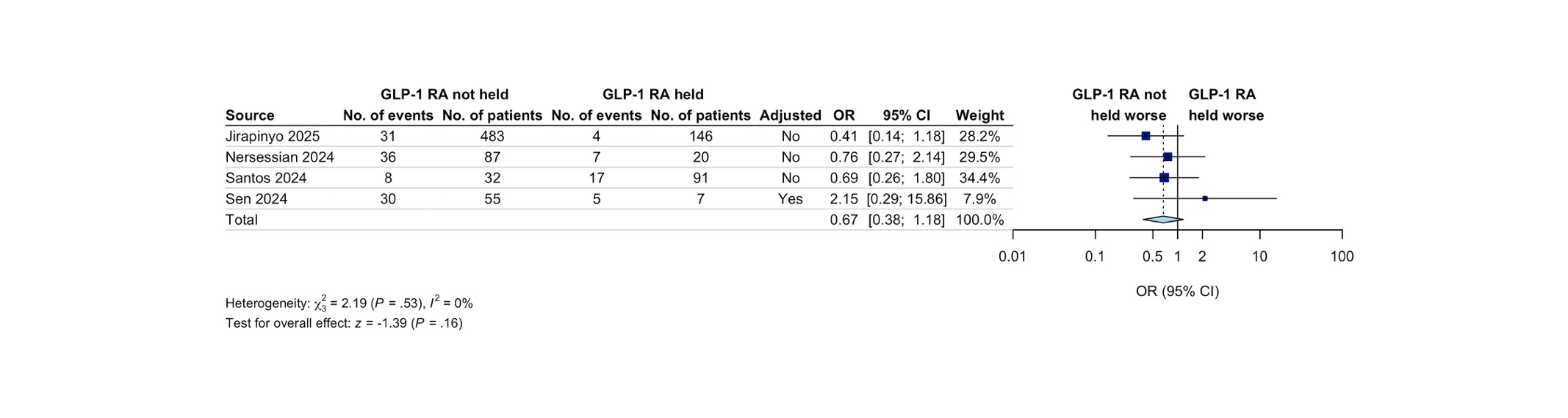

Random-effects model with restricted maximum likelihood heterogeneity variance estimator. The dark blue boxes represent individual study odds ratio, and the size of the boxes are proportional to study weight in the meta-analysis; the whiskers represent the confidence intervals; light blue diamond represents the overall pooled odds ratio and 95% CI; the dotted vertical line indicates the pooled OR. Adjusted indicates adjusted ORs. Abbreviations: GLP-1 RA, Glucagon-Like Peptide-1 Receptor Agonist.

### **Figure S14. Subgroup analysis for studies assessing withholding at least one GLP-1 RA dose prior to a procedure on residual gastric contents, comparing studies limited to patients undergoing upper endoscopy procedures to all other studies**

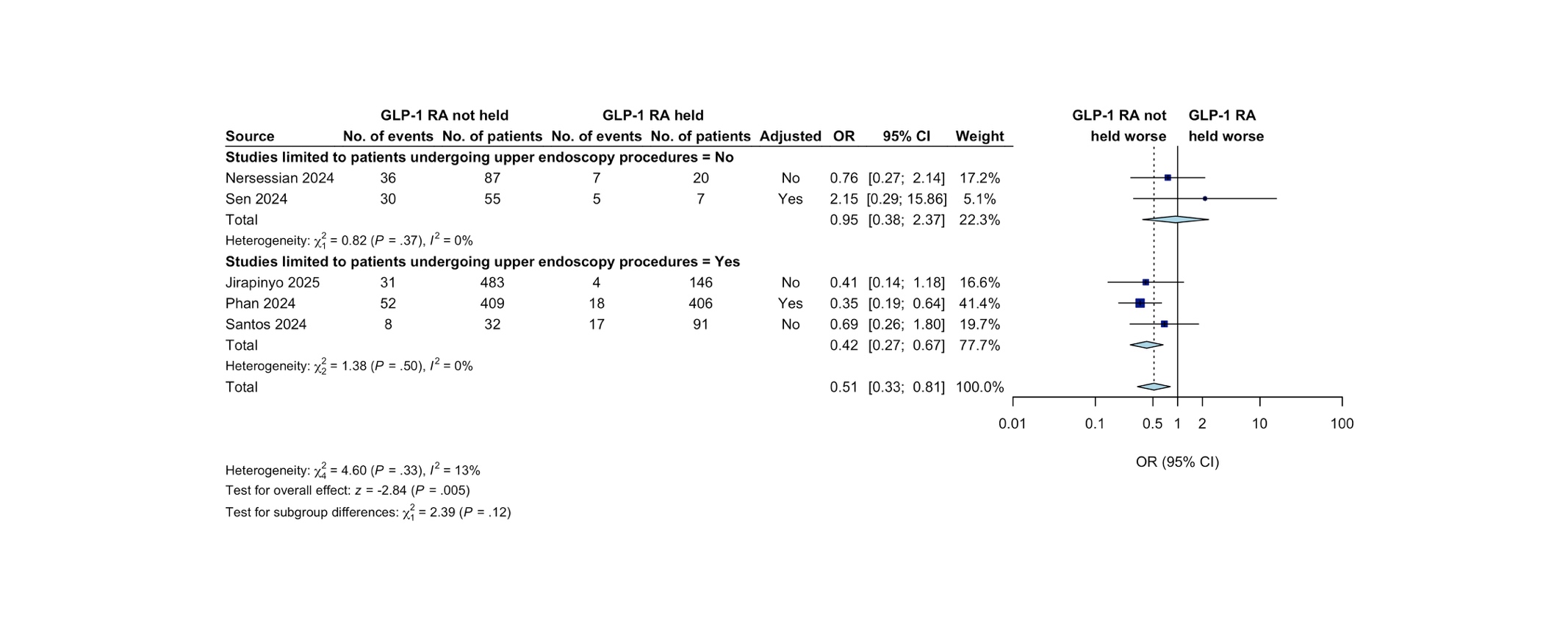

Random-effects model method: restricted maximum likelihood heterogeneity variance estimator. The dark blue boxes represent individual study odds ratio, and the size of the boxes are proportional to study weight in the meta-analysis; the whiskers represent the confidence intervals; light blue diamond represents the overall pooled odds ratio and 95% CI; the dotted vertical line indicates the pooled OR. Abbreviations: GLP-1 RA, Glucagon-Like Peptide-1 Receptor Agonist.

### **Figure S15. Subgroup analysis for studies assessing withholding at least one GLP-1 RA dose prior to a procedure on residual gastric contents, comparing studies limited to once-weekly formulations to studies not limited to once-weekly formulations.**

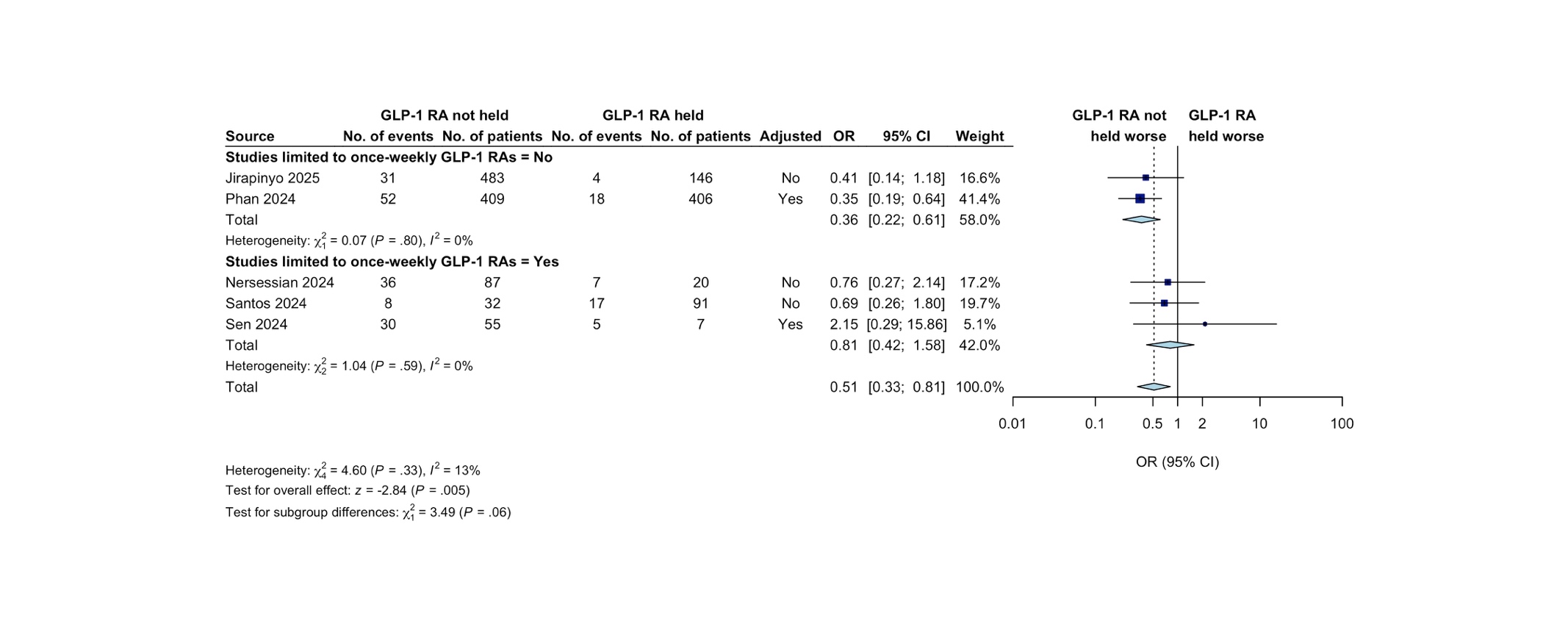

Random-effects model method: restricted maximum likelihood heterogeneity variance estimator. The dark blue boxes represent individual study odds ratio, and the size of the boxes are proportional to study weight in the meta-analysis; the whiskers represent the confidence intervals; light blue diamond represents the overall pooled odds ratio and 95% CI; the dotted vertical line indicates the pooled OR. Abbreviations: GLP-1 RA, Glucagon-Like Peptide-1 Receptor Agonist.

### **Table S11. Risk Of Bias In Non-randomised Studies - of Interventions (ROBINS-I) tool assessment**

| **Study** | **Outcome** | **Confounding** | **Participant selection** | **Classification of intervention** | **Deviations from intended intervention** | **Missing outcome data** | **Measurement of the outcome** | **Selection of the reported results** | **Overall Bias*** |
| --- | --- | --- | --- | --- | --- | --- | --- | --- | --- |
| Jirapinyo 2025^18^ | RGC | Serious | Moderate | Moderate | Low | No information | Moderate | Moderate | Serious |
| Nersessian 2024^22^ | RGC | Serious | Serious | Moderate | Low | No information | Low | Moderate | Serious |
| Phan 2024^2^ | RGC | Serious | Moderate | Moderate | Low | No information | Serious | Moderate | Serious |
| Santos 2024^9^ | RGC | Serious | Moderate | Moderate | Low | Moderate | Moderate | Moderate | Serious |
| Sen 2024^26^ | RGC | Moderate | Serious | Moderate | Low | Low | Low | Low | Serious |

Green indicates low risk of bias; Yellow indicates moderate risk of bias; Orange indicates serious risk of bias; Red indicates critical risk of bias; Grey indicates no information to form a judgement about risk of bias. Abbreviations: RGC, residual gastric contents.

* Studies were rated as low risk of bias if all domains were marked as low; Studies were rated as moderate risk of bias if all domains are either low or moderate; Studies were rated as serious risk of bias if at least one domain was marked as serious, but not at critical risk of bias in any domain; Studies were rated as critical risk of bias if at least one domain was marked as critical.

.
